# Durable Response and Improved CD8 T Cell Plasticity in Lung Cancer Patients After PD1 Blockade and JAK Inhibition

**DOI:** 10.1101/2022.11.05.22281973

**Authors:** Divij Mathew, Melina E. Marmarelis, Caitlin Foley, Joshua M. Bauml, Darwin Ye, Reem Ghinnagow, Shin Foong Ngiow, Max Klapholz, Soyeong Jun, Zhaojun Zhang, Robert Zorc, Maximillian Diehn, Wei-Ting Hwang, Nancy R. Zhang, Corey J. Langer, E. John Wherry, Andy J. Minn

**Affiliations:** Department of Systems Pharmacology and Translational Therapeutics; Department of Medicine; Department of Radiation Oncology; Department of Biostatistics, Epidemiology and Informatics; Institute for Immunology; Abramson Family Cancer Research Institute; Parker Institute for Cancer Immunotherapy; Mark Foundation Center for Immunotherapy, Immune Signaling, and Radiation, Perelman School of Medicine, University of Pennsylvania, Philadelphia, PA, USA; Department of Radiation Oncology and Stanford Cancer Institute, Stanford University School of Medicine, Stanford, CA 94305, USA; Department of Statistics, The Wharton School, University of Pennsylvania, Philadelphia, PA

**Author notes:** These authors contributed equally. Correspondence to: Andy J. Minn, MD, PhD 421 Curie Blvd BRB II/III, Room 510 Philadelphia, PA 19104.

## Abstract

Persistent inflammation including type-one interferon (IFN-I) can cause immunosuppression. We show that delayed administration of the JAK1 inhibitor itacitinib after anti-PD1 improves immune function and anti-tumor response in mice, and results in high response rates (67%) in a phase-2 clinical trial for metastatic non-small cell lung cancer with tumor PDL1≥50%. In contrast to patients with low inflammation who responded to anti-PD1, patients with elevated inflammation had poor immune and tumor responses to anti-PD1 that improved after adding itacitinib. Itacitinib promoted features of CD8 T cell plasticity and therapeutic responses of exhausted and effector-memory clonotypes. Patients with persistent IFN-I signaling refractory to itacitinib showed progressive CD8 T cell terminal differentiation and progressive disease. Thus, JAK inhibition may improve anti-PD1 efficacy by pivoting T cell differentiation dynamics.

---

The development of monoclonal antibodies to block inhibitory receptors has led to durable response to many different cancer types. Most notable is blockade of the PD1/PDL1 signaling axis to reactivate anti-tumor CD8 T cells. In non-small cell lung cancer (NSCLC), the response rate to single agent pembrolizumab (anti-PD1) is approximately 45% in the first-line metastatic setting for patients with tumor PDL1 ≥ 50%^1^. In this select group, this response rate is greater than with chemotherapy alone, making immunotherapy the current standard of care for first-line therapy of patients with metastatic NSCLC that is PDL1 ≥ 50%. Despite this improvement, approximately two-thirds of patients with NSCLC who respond to pembrolizumab will relapse^2^, making development of approaches that can improve response rates and/or augment the durability of responses an important goal in immune checkpoint blockade (ICB) therapy.

Interferon (IFN) signaling, which utilizes the Janus kinase (JAK) family, has well recognized roles in immune stimulation and promoting anti-tumor immunity. However, the role of IFN signaling in immunity is complex and IFN signaling can also have immunoregulatory effects. For example, in chronic LCMV infection, which is associated with high levels of IFN-stimulated genes (ISGs), persistent type-one interferon (IFN-I) signaling can inhibit ongoing immune responses by disrupting splenic architecture, increasing PDL1 expression, and thereby limiting viral clearance^3^. These effects were largely reversible by blocking the IFN-I receptor (IFNAR1), suggesting a therapeutic benefit to inhibiting chronic IFN signaling. Moreover, blockade a week or more post infection still showed therapeutic benefit in reducing viral burden, indicating the deleterious effect of chronic IFN signaling is reversible after initiation of chronic infection^4, 5^. IFN-I blockade also prevented antigen-specific CD8 T cells from becoming terminally exhausted and expanded the CXCR5^+^ TCF1^+^ exhausted progenitor subset^6^. Consistent with these observations, JAK inhibitors, which also inhibit IFN signaling, can restore type-two IFN (IFNG) production in LCMV derived exhausted CD8 T cells and target maladaptive IFN signaling^7^.

In cancer, which represents another disease characterized by chronic inflammation, persistent IFN signaling in cancer cells renders tumors resistant to ICB^8^. Indeed, high levels of a subset of ISGs in cancer cells is associated with immunotherapy resistance in multiple human tumors, including in NSCLC after acquired resistance to anti-PD1 blockade^2^. In CD8 T cells from patients with NSCLC and other types of cancer, high levels of ISGs are coupled to differentiation toward terminal states that include T cell exhaustion^9^. Moreover, mutations of IFN pathway genes in NSCLC patients predicts longer progression-free survival, and blocking IFN signaling in ISG-high mouse cancer models improves immune function and tumor responses to ICB^8, 10^. Thus, maladaptive or persistent IFN signaling in cancer cells and cancer-associated immune cells is linked with ICB resistance in humans and impedes the efficacy of immunotherapy in mice.

## RESULTS

### JAK inhibition to interrupt persistent IFN-I signaling improves immune function and response in mouse tumors resistant to immune checkpoint blockade

We previously demonstrated in mice that a JAK1/2 inhibitor (ruxolitinib) given after the start of ICB can re-sensitize ICB-resistant tumors from multiple cancer types^8^. To examine if the JAK1 selective inhibitor itacitinib can similarly improve ICB response, we used the Res 499 tumor model, a well-characterized ICB-resistant tumor derived from B16-F10 melanoma^11^. Administration of itacitinib seven days after the start of anti-PDL1 plus anti-CTLA4 resulted in improved tumor response (**Fig. 1A-B, S1A**). Since JAK inhibitors (JAKi) can inhibit numerous cytokine signaling pathways besides IFN, we also compared the effects of itacitinib to anti-IFNAR1. This comparison demonstrated that anti-IFNAR1 largely phenocopied itacitinib, suggesting that inhibiting IFN-I signaling is an important property of, and sufficient for, the JAKi effect. Examination of leukocytes from the tumor or spleen by flow cytometry revealed that the combination of itacitinib or anti-IFNAR1 with ICB promoted similar overall changes in leukocyte composition (**Fig. 1C, S1B**). Among the leukocytes most changed after addition of itacitinib or anti-IFNAR1 to ICB were B cells and CD8 T cells (**Fig. S1C-D**).

**Figure 1.**
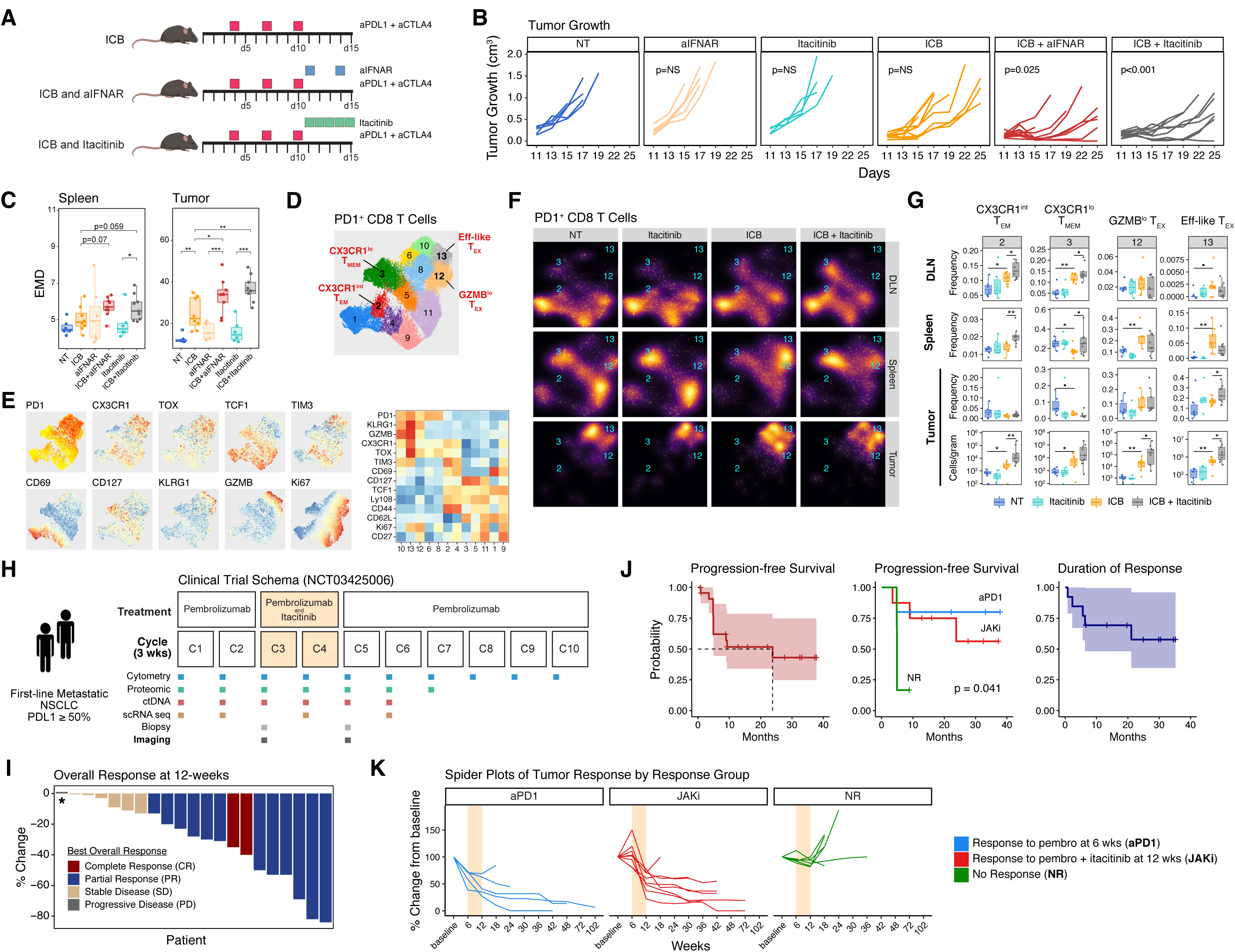
High response rates to anti-PD1 and a JAK1 inhibitor in pre-clinical studies and a phase 2 clinical trial for non-small cell lung cancer. **A)** Treatment regimen with ICB plus either itacitinib, a JAK1 inhibitor, or anti-IFNAR1 antibody for mice bearing resistant Res 499 tumors. **B-C)** Tumor growth curves **(B)** and changes in tumor and peripheral (spleen) leukocytes (day 16) measured by flow cytometry, dimensionality reduction, and Earth Mover’s Distance (EMD) **(C)**. **D-F)** PD1^+^ CD8 T cells from the tumor, draining lymph node (DLN), and spleen. Shown is a UMAP of 12 clusters **(D)**, an overlay of the expression for select markers **(E)**, and an overlay of the density distribution by treatment group and tissue **(F)**. Cluster 7 was removed due to sparsity. **G)** The proportion of the PD1^+^ CD8 T cells in the indicated tissue and absolute counts from the tumor (bottom row) for select effector, memory, and T_EX_-like clusters shown in (D). **H)** Schema of phase 2 clinical trial for pembrolizumab and delayed administration of itacitinib for first-line metastatic NSCLC with tumor PDL1 ≥ 50%. Treatment, sample collection, and response assessment by imaging are shown relative to each 3-week treatment cycle. **I)** Waterfall plot of 12-week response color-coded by best objective response. Asterisk indicates a patient who clinically progressed prior to the 12-week assessment. **J-K)** Survival curves for overall progression-free survival, progression-free survival by response group, and overall duration of response (J). Patients were categorized into three response groups: complete response (CR) or partial response (PR) after pembrolizumab but prior to itacitinib (aPD1), CR or PR only after itacitinib (JAKi), or non-responders (NR). Spider plots indicating change in tumor measurements from baseline is shown for patients in each response group (K). The window of JAKi treatment is indicated by the beige box. Significance for tumor growth was determined by a mixed-effect regression model. For pairwise comparisons, a two-sided Wilcox test or t-test was used for non-parametric or parametric data, respectively. Survival differences were determined by a log-rank test. • p<0.1, * p<0.05, ** p<0.01, *** p<0.001.

Since IFN-I can promote dysfunction of CD8 T cells^12^, we opted to subsequently narrow our focus on PD1^+^ CD8 T cells and systemically evaluate changes in the tumor, draining lymph node (dLN), and the spleen. From the flow cytometry analysis, these cells were classified into 12 clusters that differentially distributed between the three tissues (**Fig. 1D-F**). Of these clusters, clusters 2, 3, and 13 significantly increased in the dLN or tumor when JAKi was added to ICB (**Fig. 1G**). Cluster 3 resembled CX3CR1^lo^CD62L^+^CD127^+^ memory CD8 T cells (CX3CR1^lo^ T_MEM_), while cluster 2 was a CX3CR1^int^CD62L^−^ population that resembled effector-memory cells (CX3CR1^int^ T_EM_). Both CD8 T cell populations increased in frequency in the dLN and spleen after ICB and further increased with addition of JAKi. In the tumor, JAKi plus ICB increased the total number of PD1^+^ CD8 T cells (per gram of tumor) more than ICB alone (**Fig. S1E**). This increase in tumor-infiltrating CD8 T cells included the CX3CR1^int^ T_EM_ cluster 2 cells (**Fig. 1G**, bottom two rows). Cluster 13 cells also increased in frequency and cell numbers in the tumor when JAKi was added to ICB. Cluster 13 CD8 T cells had high expression of PD1 and TOX as well as CX3CR1, KLRG1, Ki67, and GZMB, consistent with a T_EX_ phenotype with effector-like features and resemblance to a previously described T_EX_ intermediate or circulatory subset^13–15^. Other T_EX_-like clusters (clusters 10 and 12) were also abundant in the tumor but did not significantly change in frequency after ICB (**Fig. 1G, S1F**). In total, these pre-clinical data confirm that itacitinib improved ICB efficacy in resistant tumors characterized by high ISGs and functioned through antagonizing IFN-I signaling to improve responses. This improved anti-tumor efficacy was accompanied by systemic increases in PD1^+^ CD8 T cell populations with features of exhaustion and in PD1^+^ CD8 T cells with effector-memory phenotypes.

### Clinical response patterns in a phase 2 clinical trial of pembrolizumab combined with a JAK1 inhibitor for first-line metastatic non-small cell lung cancer

Motivated by our pre-clinical findings, we initiated a phase two clinical trial of pembrolizumab and delayed itacitinib for treatment-naïve metastatic NSCLC with tumor PDL1 ≥ 50%. A total of 21 patients were treated and evaluated to examine the efficacy of adding itacitinib to pembrolizumab (**Fig. 1H**). Patients received two cycles of pembrolizumab and then itacitinib was added starting on day one of cycle 3 for two cycles (six weeks). Pembrolizumab was continued during concurrent itacitinib (cycles 3 and 4) and then until disease progression. Imaging was performed after the first two cycles of pembrolizumab (at week 6) and then after itacitinib (at week 12). Objective response rate (ORR) was defined as the proportion of evaluable patients with a complete response (CR) or partial response (PR) on the week 12 scan.

Clinicopathological characteristics of the 21 evaluable metastatic NSCLC patients were comparable to reported cohorts from other large academic institutions^16^ and to the pembrolizumab arm of the KEYNOTE-24 multi-country randomized trial^1^ (**Table S1**). The 12-week ORR was 62% and the best ORR with additional follow up after 12 weeks was 67% (**Fig. 1I**). After a median follow-up time of 27.6 months, the median progression-free survival (PFS) was 23.8 months (95% CI 4.9 to NA) and the median duration of response has not been reached (**Fig. 1J, S1G**). For comparison, the expected ORR from randomized studies and select U.S. academic centers is approximately 44% with a median PFS of 6.5 to 10.3 months, and the median duration of response for this patient cohort has been reported to be about 6 months^1, 16, 17^.

In addition to the 12-week response assessment, we also assessed response at 6 weeks. This analysis revealed that five patients exhibited an early (cycle 1-2) radiographic response to pembrolizumab before the addition of itacitinib. In contrast, eight patients failed to respond or had tumors that grew after initial pembrolizumab but responded at week 12 after itacitinib (cycles 3-4). Six patients remained refractory and one responder did not have a 6-week scan, precluding assessment of whether response occurred before or after addition of itacitinib. Based on these response patterns (**Fig. 1K**), we classified patients as either 6-week anti-PD1 responders (anti-PD1 responders), 12-week post-itacitinib responders (JAKi responders), or non-responders (NR). These three response groups had similar clinicopathological features (**Table S2**) but the PFS stratified by these groups was expectedly different (**Fig. 1J**, middle plot). In total, these findings suggest that the delayed administration of JAKi after anti-PD1 resulted in a higher-than-expected ORR and more durable responses in NSCLC patients with tumor PDL1 ≥ 50%. Of the responders, some patients responded early after anti-PD1 while others objectively responded only after addition of JAKi and anti-PD1.

### Differences in clinical response to anti-PD1 and JAKi are accompanied by characteristic changes in peripheral CD8 T cells

To examine how peripheral CD8 T cells changed in patients with NSCLC after treatment with anti-PD1 and delayed administration of itacitinib, we performed multi-parameter flow cytometry on CD8 T cells from peripheral blood mononuclear cells (PBMCs) (**Fig. 1H, S2A**). This analysis enabled us to examine effects resulting from initial administration of anti-PD1 (cycles 1-2), during the period of concurrent JAKi treatment (cycles 3-4), and after return to anti-PD1 monotherapy (cycles 5+). To track treatment responses, we measured Ki67 in non-naïve peripheral CD8 T cells. These data revealed a transient increase in Ki67^+^ cells from cycle 1 to 2 (**Fig. 2A**, left plot; **Fig. S2A**) corresponding to the single proliferative burst in CD8 T cells typically observed within the first two cycles of anti-PD1 in other settings^18^. After the 6-week window of concurrent itacitinib, there was a trend toward a second modest rise in Ki67^+^ CD8 T cells (start of cycle 6-7). Analysis of individual response groups shows a statistically significant increase in Ki67^+^non-naïve CD8 T cells in anti-PD1 responders after the first two cycles of anti-PD1. In contrast, JAKi responders lacked this initial Ki67 response and instead increased Ki67 at cycles 5-6 after the period of itacitinib (**Fig. 2B**, right plots). Non-responders had an inconsistent proliferative pattern throughout.

**Figure 2.**
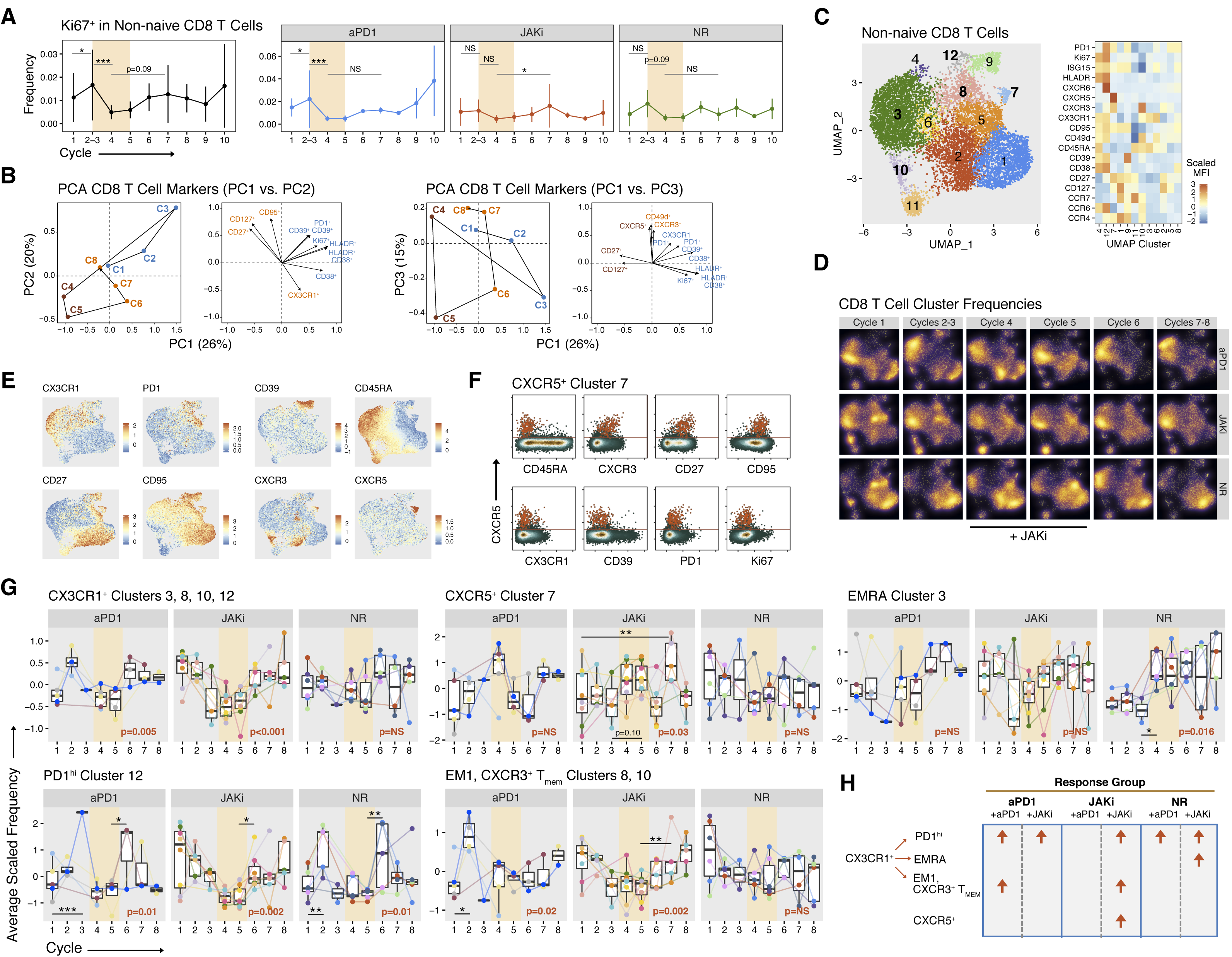
Distinct longitudinal changes in differentiation and activation of CD8 T cells associate with anti-PD1, JAK inhibition, and patient response. **A)** Percentage of non-naïve CD8 T cells positive for Ki67 for all patients (left) and stratified by response group (right plots). The start of each cycle is shown on the x-axis as well as the window of JAKi treatment (beige box). Cycles 2 and 3 are binned. **B)** Principal components (PC) analysis of manually gated flow cytometry features. The center of mass for the treatment cycles (left plot in each pair) and significant feature loadings (right plot in each pair) for the first three PCs are shown. **C)** UMAP and cluster assignment of peripheral CD8 T cells analyzed by flow cytometry (left) along with a heatmap of scaled MFI for indicated protein markers. **D)** Density distribution of cells overlaid on the CD8 T cell UMAP from (C) stratified by treatment group (rows). **E)** Scaled MFI for the indicated markers overlaid on the CD8 T cell UMAP from (C). **F**) Co-expression on non-naïve CD8 T cells between CXCR5 and each of the indicated markers, which are associated with progenitor-like CD8 T cells reported from human NSCLC patients. The red dots represent cells from the CXCR5^+^ cluster 7 from the CD8 T cell UMAP shown in (C). **G-H)** Changes in the average scaled frequency for each of the indicated CD8 T cell clusters measured at the start of the indicated treatment cycle and faceted by response group **(G)**. Individual patients are color-coded and the window of JAKi treatment is shown (beige). The main-effect p-value for significance of change across treatment cycle is indicated (red) along with p-values from select comparisons from interaction analysis. The relationship of the clusters to the main findings for each treatment group are summarized in the lower right **(H)**. For longitudinal data, significance was determined by a repeated measures ANOVA using a mixed effect model and post-hoc interaction analysis. • p<0.1, * p<0.05, ** p<0.01, *** p<0.001.

Using all manually gated features, we further assessed differences in blood CD8 T cell populations using principal component (PC) analysis where the first three PCs accounted for 61% of the variance (**Fig. S2B**). Positive PC1 loadings corresponding to cycles 1 to 3 (**Fig. 2B**, blue labels) indicated strong associations with CX3CR1, Ki67, PD1, and exhaustion markers such as CD39, consistent with known effects of anti-PD1 on CD8 T cells. Negative PC1 and/or positive PC3 loadings corresponding to cycles 4 or 5, which captures effects during JAKi (brown labels, right plots), showed involvement of CXCR5^+^ CD8 T cells and T cells with effector-memory markers (CD127, CD27). Negative PC2 or positive PC3 loadings revealed that post-JAKi cycles 6+(orange-red labels) were again associated with CD8 T cells expressing CX3CR1 and effector-memory markers (CXCR3, CD127, CD27).

To understand and corroborate this PC analysis, features from equally sampled non-naïve CD8 T cells were embedded into UMAP space for unbiased clustering (**Fig. 2C**). Density projections revealed that all three response groups differed at baseline and after therapy (**Fig. 2D**) with non-responders being most dissimilar and JAKi responders being generally less dissimilar to non-responders compared to anti-PD1 responders (**Fig. S2C**). To better define these CD8 T cell changes across time and between response groups, clusters with significant or near-significant changes in frequency across treatment cycles were selected (**Fig. S2D**) including CX3CR1^+^ clusters 3, 8, 10, and 12, and CXCR5^+^ cluster 7 (**Fig. 2E-F, Fig. 2G**). The CX3CR1^+^ clusters included a PD1^hi^CD39^+^ cluster (cluster 12) with resemblance to some T_EX_ populations, a CD45RA^+^CD27^−^CCR7^−^ EMRA population (cluster 3), a CD45RA^−^CD27^+^CCR7^−^ effector-memory (EM1) population (cluster 8), and a CXCR3^+^ early activation or memory-like (T_MEM_) cluster (cluster 10). The dynamics of the cluster 12 T_EX_-like cells significantly varied over time and differed for the three patient groups. Patients in the anti-PD1 responder and the non-responder groups had an increase in cluster 12 T_EX_-like cells during cycles 1-2. In anti-PD1 responders, this early rise in cluster 12 T_EX_-like cells was accompanied by a concurrent increase in EM1 and CXCR3^+^ T_MEM_ clusters. By contrast, in non-responders the early increase in T_EX_-like cells was followed by a progressive expansion of cluster 3 EMRA cells despite addition of JAKi. For the JAKi responder group, the early expansion of cluster 12 T_EX_-like cells was absent, mirroring the muted Ki67 response. Instead, the addition of JAKi showed a trend toward increased frequency of the CXCR5^+^ cluster 7 population during JAKi that then significantly increased post-JAKi. This change in CXCR5^+^ cluster 7 was accompanied by a post-JAKi increase in both T_EX_-like cluster 12 and the EM1 and CXCR3^+^ T_MEM_ clusters. This relatively rare CXCR5^+^ CD8 T cell population is reminiscent of stem-like CXCR5^+^ CD8 T cells that have been identified in mice^6^ and in human NSCLC patients^19^. Like these and similar progenitor-like populations^20^, the CXCR5^+^ cluster 7 cells that specifically expand during JAKi were predominantly PD1^int^, Ki67^−^, CX3CR1^−^, CXCR3^+^, CD27^+^, CD95^+^, and CD39^−^ CD8 T cells (**Fig. 2F**).

In total, these findings suggest that each clinical response group was associated with distinct changes in peripheral CD8 T cells after anti-PD1 and JAKi (**Fig. 2H**). Anti-PD1 responders exhibited early expansion of Ki67^+^PD1^hi^ T_EX_-like cells that was accompanied by a concurrent increase in effector-memory CD8 T cells. In contrast, this early increase in PD1^hi^ T_EX_-like cells in non-responders was followed by a progressive increase in EMRA cells. JAKi responders lacked an obvious increase in overall frequency of Ki67^+^PD1^hi^ cells during initial anti-PD1 treatment; however, the addition of JAKi increased CXCR5^+^ CD8 T cells that might contain progenitor-like CD8 T cells. This increase in CXCR5^+^ CD8 T cells was associated with a post-JAKi expansion of Ki67^+^PD1^hi^ T_EX_-like cells and effector and memory-like subsets.

### Evolution of CD8 T cell clonotypes links differentiation to both clinical response and effects of anti-PD1 and JAKi

To track the impact of anti-PD1 and JAKi on the differentiation of T cells, we performed single-cell RNA plus T cell receptor (TCR) sequencing (scRNA/TCR-seq) on patients (n=2-3) from each clinical response group at baseline (start of cycle 1), after anti-PD1 (start of cycle 2), during concurrent JAKi therapy (start of cycle 4), and after return to anti-PD1 monotherapy (start of cycle 6). A reference CD8 T cell map for these 32 samples was created and comprised of 13 clusters annotated by expression of gene markers, enrichment of CD8 T cell atlas gene sets^9, 21^, and trajectory analysis (**Fig. 3A-B, S3A-B**). Clusters resembling early activated EM and CM subtypes (Early.EM, Early.CM) were defined by gene sets consisting of ribosomal genes and the lncRNA *MALAT1* and by intermediate enrichment for EM and CM gene sets. Also annotated are more distinctive CM subtypes (CM.IL7R), early activation memory-like subtypes expressing *CXCR3* (CM.CXCR3), and an EM subtype (EM1). Clusters comprised of more terminally committed populations consist of *CXCL13*-expressing T_EX_ (Exh.CXCL13) and two EMRA subtypes (EMRA, EMRA.NK), one of which expressed high levels of NK receptors that have been associated with T cell dysfunction^22, 23^. Also represented are subtypes suggestive of pre- or early exhausted T cells that differ by *GZMK* and *TCF7* gene expression (Exh.GZMK, Exh.TCF7) or characterized by *BCL6* (BCL6.ISG). The final two clusters correspond to naïve cells (naïve) and a rare and poorly characterized population (unannotated). Using this annotated reference, we mapped CD8 T cells from all samples, keeping only T cells with a single TCR (comprised of a rearranged alpha and beta chain) and a minimum annotation score. Finally, to enrich for treatment-relevant T cell clonotypes, a set of filtering criteria were used to include only TCR clonotypes that expanded in response to therapy (**Fig. 3C**), resulting in 1901 distinct clonotypes from 124,534 CD8 T cells.

**Figure 3.**
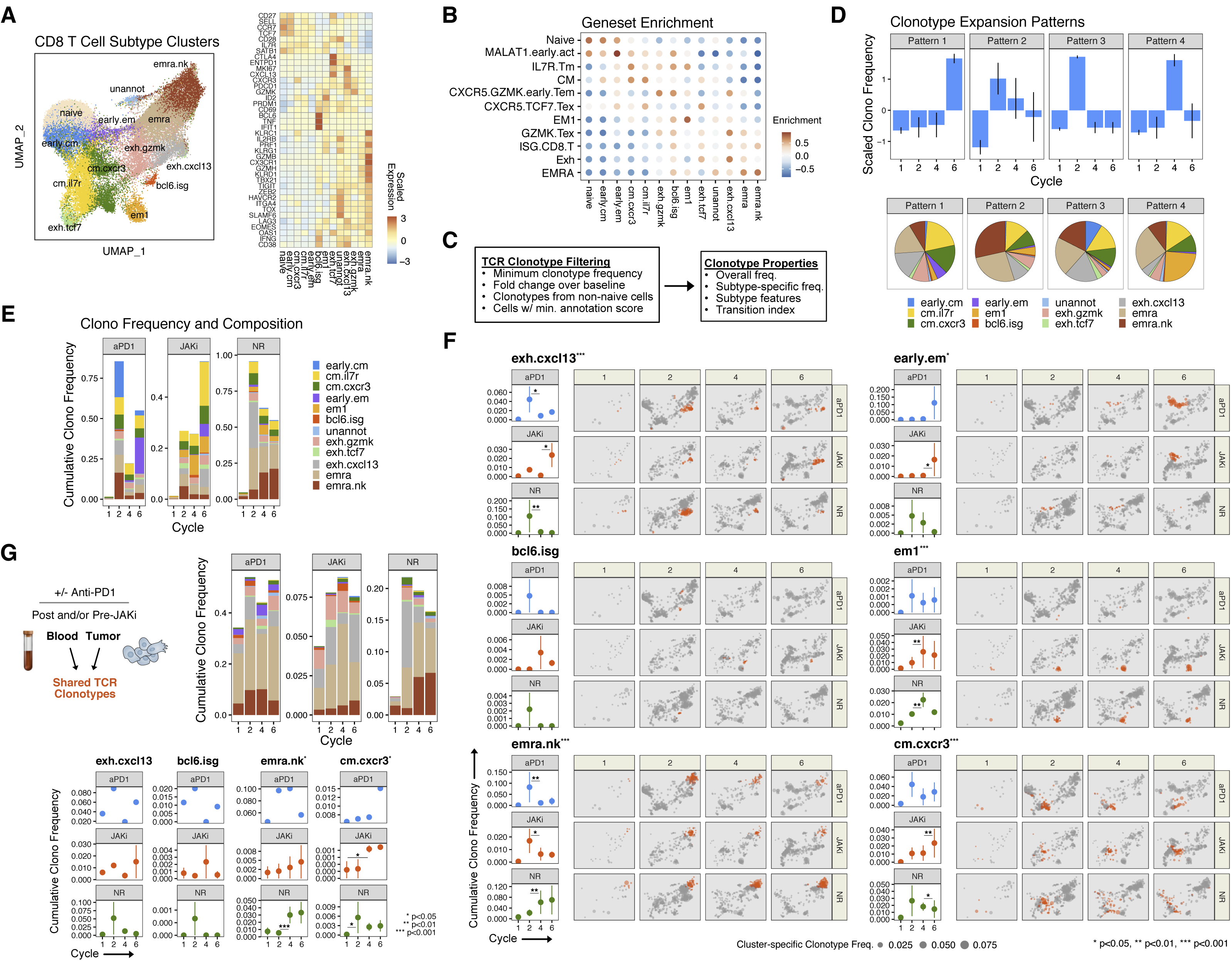
Evolution in CD8 T cell clonotypes after anti-PD1 and JAKi treatment correlate with treatment and response. **A-C)** Peripheral CD8 T cells from the start of cycles 1, 2, 4, and 6 were interrogated by scRNA/TCR-seq. UMAP clusters representing CD8 T cell subtypes (left) and heat map of scaled expression values for select marker genes (right) are shown **(A)**, as well as GSVA enrichment scores for gene sets from the indicated human CD8 T cell subtype **(B)**. CD8 T cells that meet the indicated filtering criteria were used to compile and characterize blood-expanded clonotypes (T cells sharing the same T cell receptor) for further analysis **(C)**. **D)** Patterns of clonotype expansion at the start of the indicated treatment cycle (top) and their subtype composition (bottom). Shown are scaled average frequencies and standard deviation for each cycle from analysis of all clonotypes. **E)** Cumulative subtype-specific frequency of clonotypes across treatment cycles for patients pooled by treatment group. **F)** Clonotypes belonging to the indicated subtype (red) or to other subtypes (grey) are overlaid on the CD8 T cell UMAP from (A). The size of the dots is proportional to the subtype-specific frequency. For each response group and treatment cycle, pooled patient data are displayed on the UMAP, while the mean and standard deviation calculated using individual patients are shown in the left-hand margins. Statistical significance for differences across treatment cycles is indicated by asterisks in the plot title, while significance from interaction analysis is shown in the plots in left-hand margins. Cycle 1 vs. 2 p-values are not considered due to filtering (see Methods). **G)** Cumulative subtype-specific frequency of clonotypes shared by T cells from the tumor and the blood across treatment cycles and response groups (top). The mean and standard deviation are also shown (bottom) with significance determined as in (F). For longitudinal data, significance was determined by a repeated measures ANOVA using a mixed effect model and post-hoc interaction analysis. • p<0.1, * p<0.05, ** p<0.01, *** p<0.001.

To understand how T cell clonotypes responded to anti-PD1 and JAKi, we first grouped the clonotypes into expansion patterns and assessed the overall distribution of T cell subtypes associated with each pattern (**Fig. 3D, S3C-D**). This approach revealed four major patterns. Approximately 20% of clonotypes that expanded at the start of cycle 2 were Exh.CXCL13 T_EX_ cells (pattern 3), consistent with effects of anti-PD1. However, a similarly large proportion of Exh.CXCL13 T_EX_ cells along with effector and memory-like CD8 T cells were also observed among clonotypes that increased post-JAKi (pattern 1). Clonotypes that expanded during JAKi (pattern 4) also contained a large proportion of effector and memory-like CD8 T cells, while clonotypes that were elevated throughout therapy (pattern 2) contained a large fraction of EMRA.NK CD8 T cells. When examined by response group (**Fig. 3E-F**), both anti-PD1 responders and non-responders had the largest proportion of Exh.CXCL13 clonotypes at the start of cycle 2 of anti-PD1 (**Fig. 3F**, upper left). In non-responders, but not in the other groups, this was followed by a progressive increase in EMRA.NK CD8 T cell clonotypes during and after JAKi (**Fig. 3F**, lower left). In JAKi responders, EM1 CD8 T cell clonotypes expanded on JAKi concurrent with the appearance of BCL6.ISG clonotypes (**Fig. 3F**, middle). This was followed by an increase in Exh.CXCL13, Early.EM, and CM.CXCR3 CD8 T cell clonotypes post-JAKi (**Fig. 3F**, upper left; upper and lower right).

Besides clonotypes in the blood that expand after anti-PD1, clonotypes shared between CD8 T cells from tumor and blood can also enrich for tumor-relevant CD8 T cells. Despite a paucity of viable cells from most tumor biopsies due to high clinical response rates, we obtained enough tumor material from some patients for TCR-sequencing. This analysis identified 300 clonotypes out of 4072 found in tumors that were also found in blood CD8 T cells. Shared tumor-blood clonotypes were enriched for Exh.GZMK, Exh.CXCL13, EMRA, and EMRA.NK subtypes (**Fig. 3G**), likely due to the higher representation of these CD8 T cells in the tumor. TCR clonotypes comprised of the BCL6.ISG cluster (color-coded in bright red) were also enriched preferentially in the anti-PD1 and JAKi responders (p<0.001, log-linear model). Despite the small number of shared clonotypes for analysis, several key time-dependent changes found from the analysis of blood-expanded clonotypes were confirmed (**Fig. 3G**, bottom plots). For JAKi responders, there was an increase in the blood of CM.CXCR3 tumor-blood clonotypes after addition of JAKi. For non-responders, JAKi increased the frequency of EMRA.NK tumor-blood clonotypes.

Together, these findings corroborate and elaborate on our flow cytometry results (**Fig. 2G**). Specifically, in anti-PD1 responders there was an early expansion of clonotypes comprised of T_EX_-like cells, but in JAKi responders this early T_EX_-like clonotype expansion was muted. Rather, JAKi responders displayed an increase in rare BCL6.ISG clonotypes after addition of JAKi, which was similar to the increase in CXCR5^+^ cells observed by flow cytometry. The post-JAKi increase in BCL6.ISG clonotypes occurred concurrently with or was followed by expansion of both T_EX_-like clonotypes and clonotypes comprised of other CD8 T cell subtypes such as CM.CXCR3 and EM1. Non-responders, in contrast, were characterized by expansion of NK-like CD8 T cell EMRA clonotypes.

### Progenitor-like CD8 T cells link JAK inhibition to increased features of developmental plasticity

Our analyses from mice and humans collectively suggest that JAKi might affect progenitor-like CD8 T cells to promote developmental plasticity after ICB. Therefore, to investigate how developmental fates of CD8 T cells might change with JAKi and ICB, we employed a previously described pairwise transition index (pTrans-index) that measures the degree of TCR sharing between two T cell states and hence implies developmental relatedness^24^. As illustration, overlaying pTrans-index values for the rare BCL6.ISG subtype onto a UMAP of blood-expanded clonotypes summarizes how clonotypes from BCL6.ISG and other subtypes are developmentally related by treatment and response group (**Fig. 4A**). Here, BCL6.ISG clonotypes in the JAKi responder group predominantly shared TCRs with multiple effector and memory CD8 T cell clonotypes during JAKi therapy (cycle 4, middle row) but then largely shared TCRs with exhausted and non-exhausted CD8 T cell clonotypes after JAKi (cycle 6, middle row). The dynamics of the changes in TCR sharing varied by subtypes predicted to reside along different points in the developmental trajectory of CD8 T cells (**Fig. S4A-C, S3A**).

**Figure 4.**
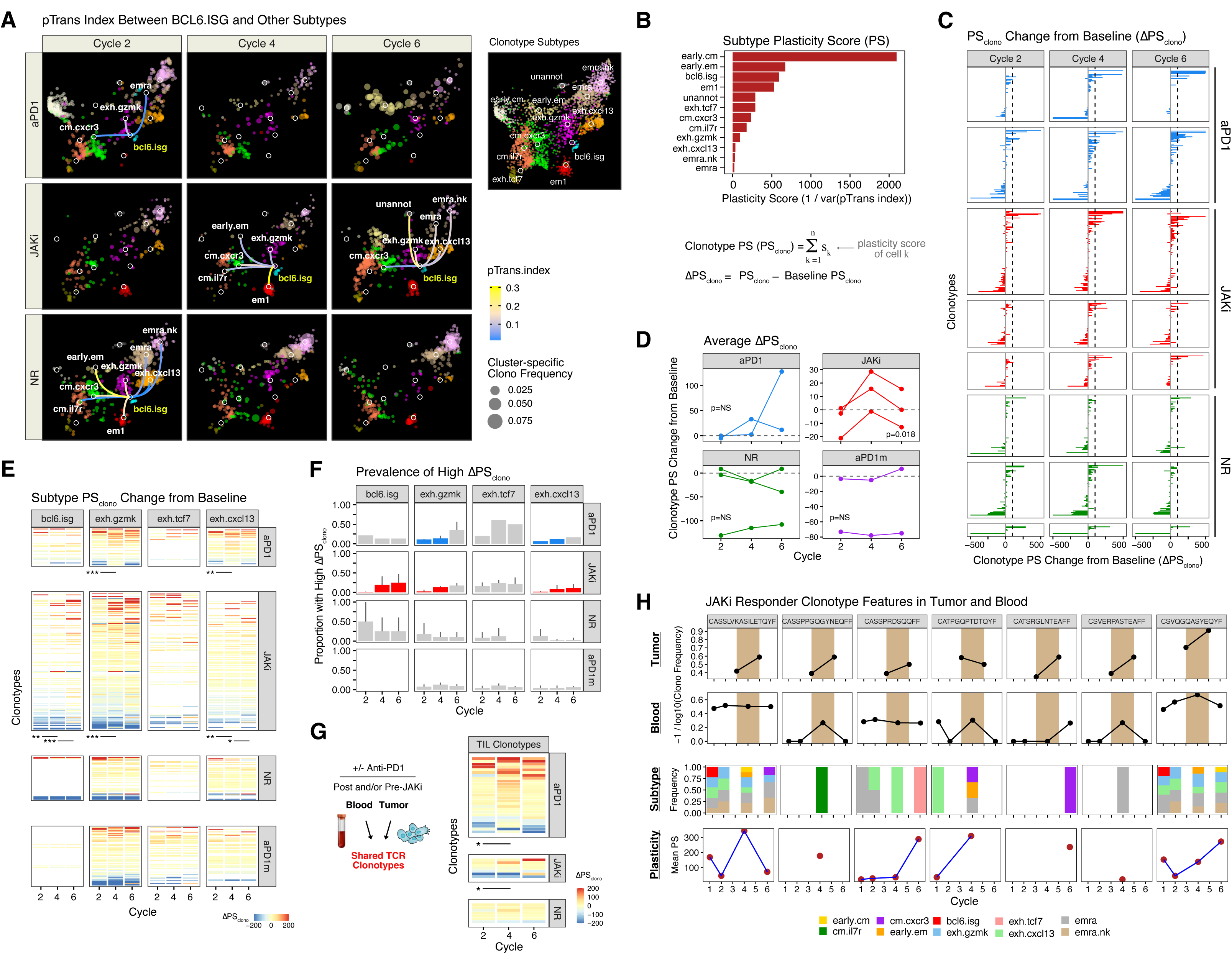
Alterations in CD8 T cell clonotype plasticity and differentiation are associated with response after addition of JAKi to anti-PD1. **A)** Blood-expanded CD8 T cell clonotypes belonging to the indicated subtype (color-coded) overlaid on the CD8 T cell UMAP (Fig. 3A) and faceted by treatment cycle and response group. The pairwise transition-index (pTrans-index) between the BCL6.ISG subtype and other subtypes are shown as lines between nodes and color-coded by the pTrans-index value (higher scores indicate greater TCR sharing). Subtype-specific clonotype frequency is represented by dot size. **B-D)** Plasticity scores (PS), derived from pTrans-indices, for each CD8 T cell subtype **(B)** were used to determine a clonotype PS (PS_clono_) and a ΔPS_clono_, which is the difference between PS_clono_ and the PS_clono_ at baseline. The ΔPS_clono_ values for expanded clonotypes from each patient in the indicated response group across treatment cycles are shown **(C)**. The mean **(D)** of the ΔPS_clono_ distributions are compared between response groups, and includes patients treated with anti-PD1 without JAKi (aPDm). Positive ΔPS_clono_ represents an altered clonotype subtype composition resulting from an increased plasticity. **E-G)** ΔPS_clono_ for clonotypes shared with each of the indicated subtypes **(E)** and the proportion of clonotypes with high ΔPS_clono_ (> 100) **(F)**. The significance for an increase in ΔPS_clono_ for paired clonotypes between the indicated cycles is shown beneath the faceted heatmaps in (E), and comparisons that are not statistically significant are indicated by grey bars in (F). Patients from an independent study treated with only anti-PD1 are also included (aPD1m). **G)** ΔPS_clono_ for clonotypes shared in tumor and blood. **H)** Clonotypes shared by CD8 T cells from blood, pre-JAKi tumor, and post-JAKi tumor from a JAKi responder. The frequency in the tumor and blood (top two rows), subtype composition (third row), and PS_clono_ (bottom row) are shown. The JAKi treatment window is shown by the beige box. For longitudinal data, significance was determined by a repeated measures ANOVA using a mixed effect model and post-hoc interaction analysis. • p<0.1, * p<0.05, ** p<0.01, *** p<0.001.

Using the pTrans-index, we derived a plasticity score (PS), whereby high PS indicates relatively even TCR sharing between a T cell subtype and multiple T cell states (as in the case of BCL6.ISG), while low PS represents strong sharing that is restricted between a subtype and only a few other states (**Fig. 4B, S4D**). By extension, the clonotype PS (PS_clono_) is based on the subtype composition and is the weighted average of the PS values of the component subtypes. For each expanded clonotype from non-naïve CD8 T cells, we then determined the change in PS_clono_ at the start of cycles 2, 4, and 6 compared to baseline (ΔPS_clono_) (**Fig. 4C**). This analysis revealed that although positive and negative changes in ΔPS_clono_ occurred across cycles, patients in the JAKi responder group had an increasing bias for multiple clonotypes with large positive shifts in ΔPS_clono_ during JAKi (cycle 4 vs. 2, red bars), which is indicative of having further increased their plasticity from baseline. This change was quantitated by a higher positive skew in the ΔPS_clono_ distribution and a greater proportion of clonotypes with a large positive change (> 100) in ΔPS_clono_ (**Fig. 4D, S4E**). Compared to anti-PD1 responders and non-responders, both metrics were consistently higher for JAKi responders during cycle 4 of JAKi treatment. Moreover, in a separate cohort of NSCLC patients not part of the trial and treated with only anti-PD1 without JAKi, the ΔPS_clono_ values remain predominantly negative or near-zero across similar therapy cycles, resembling the pattern observed in non-responders (**Fig. 4D**, purple vs green lines). Thus, these results suggest that the addition of JAKi to anti-PD1 altered clonotype composition to include less committed CD8 T cell subtypes, an effect that was particularly pronounced in patients who responded to ICB after addition of JAKi.

We next sought to examine which CD8 T cell subtypes might contribute to JAKi-related changes in clonotype plasticity. Flow cytometry data pointed toward a CXCR5^+^ progenitor-like population. Such progenitors have been reported to co-express CXCR5 with BCL6^6, 25^ suggesting possible overlap with the BCL6.ISG subtype that, like the CXCR5^+^ population, increased with itacitinib in patients from the JAKi responding group (**Fig. 2G, 3E-F**). However, the extent to which progenitor-like CD8 T cells across species and different assays represent the same population is unclear. For example, low abundance of *CXCR5* transcripts prevented assessment of *CXCR5* expression by the BCL6.ISG population. Nonetheless, human atlas data for tumor-associated T cells have revealed that CXCR5^+^ CD8 T cells are split between *GZMK* early T_EM_ and *TCF7* T_EX_ populations^9^, two subtypes described to have developmental relationships with multiple subtypes based on TCR sharing. Genes from each of these CXCR5^+^ CD8 T cell populations are enriched in BCL6.ISG but also in the Exh.GZMK and Exh.TCF7 subtypes, and to a lesser degree, the Exh.CXCL13 subtype (**Fig. 3B**). Additionally, these candidate progenitors were also moderately to strongly enriched in ISGs (ISG.CD8.T gene set), which is characteristic of progenitor CD8 T cells^6^. Therefore, we examined the clonotypes shared with each of these subtypes to assess how the ΔPS_clono_ changed over treatment (**Fig. 4E**). In anti-PD1 and/or JAKi responders, the addition of JAKi led to a significant increase in ΔPS_clono_ for clonotypes from each subtype (cycle 2 vs. 4) except for those shared with Exh.TCF7. However, only in JAKi responders did ΔPS_clono_ continue to increase after JAKi (cycles 4 vs. 6), namely for the BCL6.ISG and Exh.CXCL13 clonotypes. Of the clonotypes and timepoints showing significant increases in ΔPS_clono_ (**Fig. 4F**, non-grey bars), BCL6.ISG clonotypes in particular had a high proportion with large positive changes in ΔPS_clono_. In contrast, clonotypes from non-responders and patients treated with anti-PD1 without JAKi (aPD1m) did not exhibit significant changes in ΔPS_clono_ across treatment cycles. Together, these data suggest that addition of JAKi to ICB may impact progenitor-like CD8 T cells, particularly those belonging to BCL6.ISG, to promote a more balanced differentiation toward exhausted and non-exhausted states. In JAKi responders, this effect was progressive and continued after the window of JAKi treatment, while in non-responders or patients treated with anti-PD1 alone, the effect appeared largely absent.

### Tumor-blood CD8 T cell clonotypes develop subtype composition reflecting increased developmental plasticity after JAKi

To further examine how JAKi could potentially influence the plasticity of tumor-relevant CD8 T cells in the context of PD-1 blockade, we next examined JAKi-related changes in CD8 T cell clonotypes shared by the tumor and blood. Despite high response rates resulting in limited biopsy material, a small number of shared clonotypes were available for analysis. These blood-tumor shared clonotypes demonstrated a similar pattern of greater ΔPS_clono_ after addition of JAKi in anti-PD1 and JAKi responders (**Fig. 4G**, cycles 2 vs. 4). In one JAKi responder, enough on-treatment biopsy material from both pre-JAKi (start of cycle 3) and post-JAKi (start of cycle 6) tumors were available for TCR-sequencing, enabling analysis of tumor-blood clonotypes both before and after addition of JAKi to anti-PD1 (**Fig. 4H**). For these seven tumor-blood clonotypes, six increased in frequency in the tumor from pre-JAKi to post-JAKi. Of four clonotypes detectable in the peripheral blood at multiple time points (left to right, clonotypes 1, 3, 4, 7), all increased frequency either during JAKi (start of cycle 4) or after completion of JAKi (start of cycle 6). Furthermore, these four clonotypes concurrently increased their PS_clono_, with three changing their composition during or after JAKi to include effector or memory CD8 T cell subtypes. In fact, although all clonotypes detectable in the blood just before JAKi were solely comprised of T_EX_ or EMRA-like subtypes, five out of the seven total clonotypes examined altered their composition after addition of JAKi to include effector or memory CD8 T cell subtypes. Thus, these observations detail the consequences of potentially increased CD8 T cell plasticity in a patient who responded to anti-PD1 after addition of JAKi – namely, the development of tumor-relevant clonotypes that include effector and memory CD8 T cell subtypes.

### Persistent inflammation and discordant cytokine signaling in immune cells are associated with tumors that fail to respond to anti-PD1 and JAKi

In contrast to patients who respond to anti-PD1 and itacitinib, non-responders showed marginal changes in features of CD8 T cell plasticity that mimicked findings in patients treated with anti-PD1 alone (**Fig. 4E-G**). These observations suggested that non-responders may be at least partly refractory to JAK inhibition. To investigate this notion and evaluate how inflammatory signaling correlated with clinical response, we profiled approximately 92 serum proteins and jointly analyzed pathway-related transcriptional changes in PBMCs. We first analyzed serum proteins and cytokines (**Fig. S5A**) that significantly changed with time and grouped them by temporal expression patterns. Four distinct patterns were identified, with some proteins/cytokines belonging to multiple patterns (**Fig. 5A**, left and middle plots). Anti-PD1 responders and JAKi responders were both characterized by several proteins that markedly increased and peaked after anti-PD1 at the start of cycle 2 and diminished during JAKi (pattern 4). This pattern included CXCL9 and CXCL10, well-known ISGs and ligands for CXCR3. JAKi responders were additionally characterized by proteins that were elevated at baseline, more modestly peaked after anti-PD1, and decreased after addition of JAKi (pattern 3). This pattern also included immunosuppressive cytokines (labeled red vs. blue) such as IL10, CSF1, and IL6 that, although elevated at baseline and increased during anti-PD1, were decreased by JAKi. Non-responders had elevated baseline expression of a large set of proteins that included suppressive cytokines, such as IL10, IL6, and CSF1, and this set of proteins were essentially unaffected by therapy in these patients (pattern 2). This pattern associated with non-responders was enriched in pathways linked to chronic inflammation and disease, such as IL23 and IL27 (**Fig. 5A**, plots in right margin).

**Figure 5.**
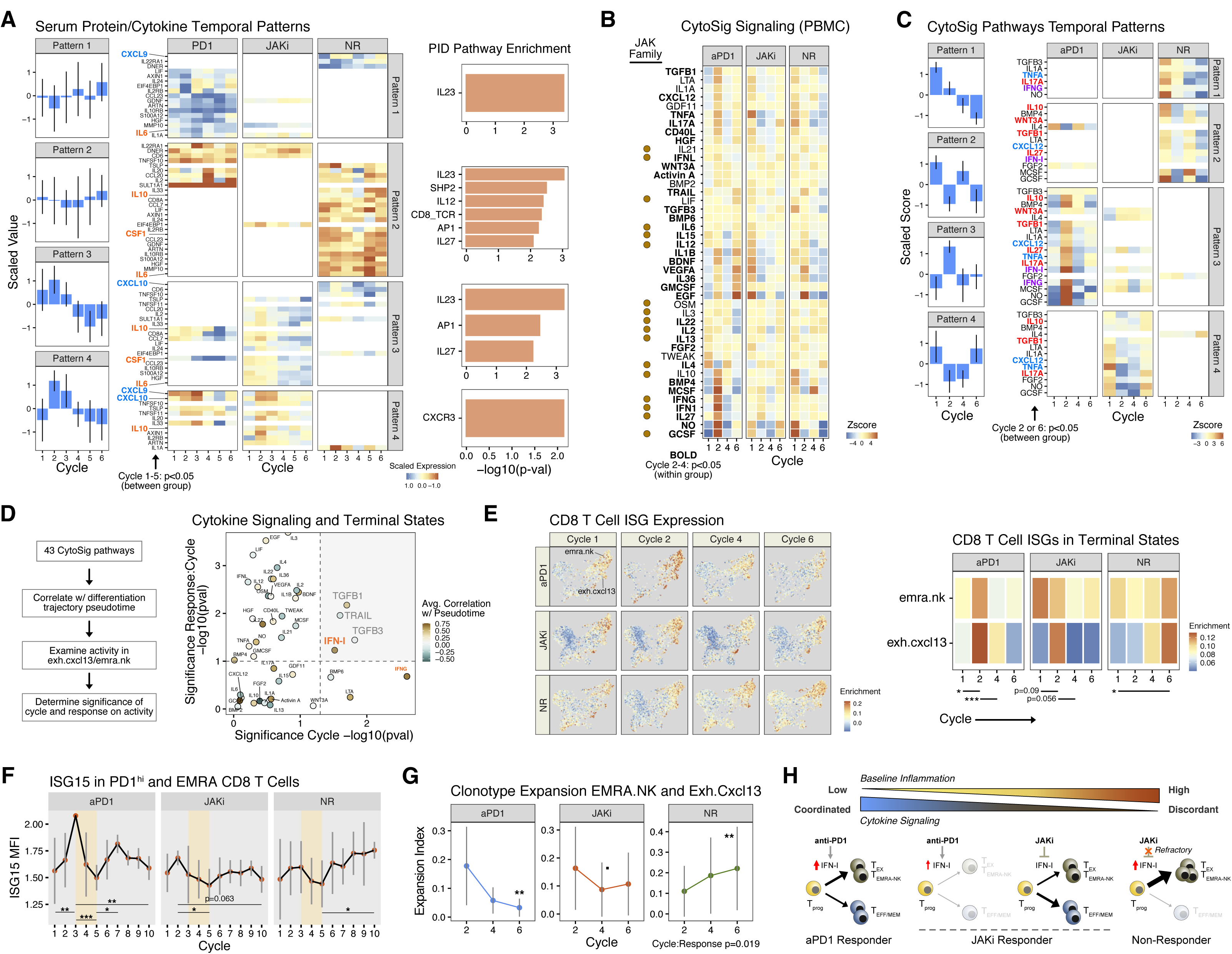
Persistent inflammation and refractoriness to JAKi are associated with terminal CD8 T cell differentiation and therapy failure. **A)** Temporal expression patterns (left) and serum levels (middle heat maps) of proteins and cytokines that significantly differ by response group. Select cytokines that are typically stimulatory (blue) or inhibitory (red) are highlighted. Pathway analysis of the proteins and cytokines that belong to each temporal pattern are also shown (right margin). **B)** Activity of cytokine signaling pathways inferred by CytoSig from PBMCs. Pathways that significantly differ at the start of cycles 2 or 4 within a response group are in bold and those that are regulated by the JAK family are highlighted (brown dot). **C)** Temporal patterns of cytokine activity that significantly differ between response groups either pre-JAKi (cycle 2 start) or post-JAKi (cycle 6 start). Select pathways that are typically stimulatory (blue) or inhibitory (red) are highlighted along with IFNG and IFN-I (purple). **D)** Cytokine pathway activity associated with CD8 T cell differentiation, treatment cycle, and response. Correlation of pathway activity with pseudotime from trajectory analysis using all CD8 T cell subtypes are color-coded. The significance of changes in pathway activity in EMRA.NK and Exh.CXCL13 subtypes across treatment cycles (main effect) is shown on the x-axis, and an interaction between this and response group (interaction effect) is shown on the y-axis. Dotted lines represent significance levels (p=0.05 for main effect, p=0.10 for interaction effect) and grey upper-right quadrant show pathways that significantly differ by main and interaction effects. **E)** Enrichment in CD8 T cell ISGs across all subtypes overlaid on a CD8 T cell UMAP (left) and specifically for EMRA.NK and Exh.CXCL13 subtypes shown as a heat map(right). **F)** ISG15 expression in PD1^hi^ and EMRA-like CD8 T cells. The MFI from flow cytometry for clusters 12 and 3 from Fig. 2C are shown. **G)** Clonotype expansion index for EMRA.NK and Exh.CXCL13 subtypes. Values measure TCR clonality of the CD8 T cells within the subtypes. **H)** Model summarizing relationship between inflammation, cytokine signaling in immune cells after anti-PD1, the impact of IFN-I on CD8 T cell differentiation toward either terminal or less committed states, and consequence of JAK inhibition. For longitudinal data, significance was determined by a repeated measures ANOVA using a mixed effect model and post-hoc interaction analysis. • p<0.1, * p<0.05, ** p<0.01, *** p<0.001.

To complement the serum cytokine/protein studies with the activation status of cytokine signaling pathways in immune cells, we next applied CytoSig^26^, a method to assess 43 signaling pathways based on transcriptional changes, to scRNA-seq data from PBMCs. We first determined the overall impact of JAKi on PBMCs by analyzing which pathway activity scores significantly changed after addition of JAKi (cycle 2 vs. 4). This analysis revealed that the majority of signaling pathways known to utilize JAKs (JAK1, 2, 3) were significantly altered by JAKi (**Fig. 5B**, bold and highlighted by brown dot). Next, we focused on signaling pathways that differed before or after JAKi (cycle 2 or 6) between response groups and then categorized these pathways by temporal patterns (**Fig. 5C**). Here, each response group was predominantly defined by one of four temporal patterns. Anti-PD1 responders were associated with pattern 3, which included IFN signaling. Pattern 3 displayed low baseline activity, strong induction at the beginning of cycle 2, and then suppression by JAKi. JAKi responders also displayed pattern 3 but had weaker induction of signaling activity at cycle 2 compared to anti-PD1 responders. JAKi responders were also associated with pattern 4 characterized by pathways with more elevated baseline activity and no induction or even suppression after anti-PD1. At cycle 6 post-JAKi, pattern 4 cytokine pathways appeared to “reset” in JAKi responders by increasing in activity after patients returned to anti-PD1 monotherapy. Non-responders were associated with patterns 1 and 2 that showed high relative baseline activity. Moreover, pattern 2 cytokine pathways showed discordant signaling whereby there was induction during JAKi at cycle 4 rather than with anti-PD1 at cycles 2 and 6. Regardless of the temporal pattern though, changes in typically stimulatory pathways (blue labels) were accompanied by pathways that are often suppressive, such as TGFB, WNT3A, IL10, IL17, and IL27 (red labels).

In total, these data suggest that early response after anti-PD1 was associated with low baseline inflammation followed by induction of coordinated cytokine signaling. In JAKi responders, a more modest and limited increase in cytokine signaling occurred after anti-PD1 possibly due to elevated baseline inflammation. In both groups, addition of JAKi diminished activity of stimulatory and inhibitory pathways and may improve subsequent responsiveness. In contrast, treatment failure was associated with high and persistent inflammatory features accompanied by discordant cytokine signaling, irrespective of anti-PD1 or JAKi treatment.

### Persistent IFN signaling in CD8 T cells recalcitrant to JAKi is associated with progressive expansion of terminal subtypes and treatment failure

The association between clinical response after JAKi with enhanced features of CD8 T cell plasticity was consistent with IFN-I promoting CD8 T cell progenitor differentiation toward dysfunctional and terminal states. However, although our pre-clinical studies showing that anti-IFNAR1 can phenocopy itacitinib (**Fig. 1B**) argue that IFN-I has a prominent role in these JAKi effects, many inflammatory signaling pathways and immune cell types are affected by itacitinib. Therefore, we systematically analyzed the extent to which CD8 T cells and IFN signaling contributed to differences between clinical response groups. For this purpose, we first determined which cytokine pathways within general immune cell types significantly differed between response groups. This analysis revealed that out of 45 pathway-immune cell combinations that differed between response groups and across treatment cycles, CD8 T cells were associated with 21 and represent the largest contributor out of all major immune cell types (**Fig. S5B**). Many of these differences involved an increase in IFNG and/or IFN-I signaling in CD8 T cells from anti-PD1 responders compared to the other two groups at the start of cycle 2 during anti-PD1. Thus, CD8 T cells were a major source of variation in IFN signaling that correlated with clinical response.

We next focused on whether persistent IFN signaling and its modulation by JAKi could influence the evolution of CD8 T cell differentiation. Correlating the activity of 43 signaling pathways with pseudotime values from trajectory analysis revealed that IFN-I and IFNG were among the top four most strongly correlated pathways (**Fig. 5D**, brown hued dots; **Fig. S5C**), connecting increases in IFN signaling with more committed CD8 T cell differentiation states such as EMRA.NK and Exh.CXCL13. Restricting analysis to these two committed subtypes revealed IFN-I signaling as among the few pathways that significantly varied across treatment cycle (**Fig. 5D**, main effect, x-axis) where this variation differed by the response group (**Fig. 5D**, interaction effect, y-axis). Accordingly, at the start of cycle 2 of anti-PD1 treatment, ISGs were most strongly induced in both EMRA.NK and Exh.CXCL13 CD8 T cell subtypes in anti-PD1 responders, blunted and inconsistently increased in JAKi responders, and remained predominantly unchanged in non-responders (**Fig. 5E**). Notably, only in non-responders did JAKi fail to decrease ISG expression. These differences in ISGs were corroborated by flow cytometry for ISG15 expression in PD1^hi^ and EMRA-like CD8 T cells whereby only in non-responders did ISG15 fail to decrease immediately post-JAKi from its pre-JAKi peak (**Fig. 5F**). The persistently elevated ISGs in Exh.CXCL13 and EMRA.NK CD8 T cell subtypes correlated with increased TCR clonality in these more terminally committed states (**Fig. 5G**) and with greater frequency of EMRA-like and PD1^hi^ CD8 T cells observed by flow cytometry after addition of JAKi (**Fig. 2G**). Moreover, the elevated ISG expression in such terminal CD8 T cells was associated with hypo-responsiveness to direct IFN-I stimulation *in vitro* compared to less committed subsets (**Fig. S5D**), a phenomenon found in situations of chronic inflammation^27^ that may enable T cells to accumulate despite anti-proliferative effects of IFN. Thus, persistent IFN-I signaling and refractoriness to resetting this signaling by JAKi may play a prominent role in promoting differentiation of CD8 T cells toward terminal and dysfunction states and in therapy failure.

## DISCUSSION

The opposing functions of IFN rely on a temporal component which activates immune responses early but inhibits the response with time, especially in the setting of chronic inflammation. As shown here and in other pre-clinical studies, blocking the inhibitory function of IFNs to improve anti-tumor immunity is achievable in mice with delayed administration of JAK inhibitors or anti-IFNAR1 antibodies. We now extend this concept to humans and demonstrate that in a phase 2 clinical trial for patients with metastatic NSCLC that is tumor PDL1 ≥ 50% this therapeutic strategy in combination with anti-PD1 is feasible. Although proof of improved clinical efficacy from JAKi addition will require larger prospective randomized studies, the high response rates (67%) and long progression-free survival (23.8 months) observed in our study encourages further clinical investigation. Additional key questions for future clinical studies include investigating other settings amenable to improvement with JAKi (e.g., NSCLC with low PDL1, other cancer types, relapsed setting) and understanding parameters critical for the ability of JAKi to complement immunotherapy (e.g., duration of treatment, JAKi selectivity).

Among potential key parameters that may impact whether JAKi can improve ICB are levels and likely duration of baseline inflammation and how multiple cytokine signaling pathways, which include stimulatory and inhibitory pathways, are coordinated (**Fig. 5H**). The cellular consequences of chronic IFN and other cytokine pathways may involve changes in signaling thresholds, altered balance between positive effectors and negative regulators, and inflexible epigenetic states in key cell types such as tumor-specific CD8 T cells, other immune cells, and cancer cells that contribute to long-lasting changes in signaling behavior. Indeed, immune cells from patients that did not respond in our trial appeared overtly refractory to JAKi and may have displayed at least some of these features. A deeper understanding of how chronic signaling impacts cell behavior will provide insight into opposing context-dependent differences in JAK signaling in ICB response^28^, and potential therapeutic efficacy of strategies that “reset” signaling pathways important in immune function.

Like in mouse models of chronic infection, longitudinal examination of CD8 T cells before and after JAKi from the patients from our trial support a role for IFN-I signaling in promoting T cell differentiation, including the development of CD8 T cell exhaustion. Our findings suggest that CD8 T cells marked by CXCR5, BCL6, and ISGs might enrich for progenitor populations responsible for generating more committed subtypes through IFN-I signaling and hence targeted by JAKi treatment, a notion also corroborated by recent mouse studies^29, 30^. Conversely, recent evidence suggests that cancer cells may co-opt IFN-I signaling to drive plasticity rather than differentiation^31^. Genome-wide genetic screens in several mouse cancer models have additionally confirmed how IFN signaling in cancer cells can promote ICB resistance^32^, consistent with elevated levels of ISGs in tumors from patients with NSCLC who have relapsed after anti-PD1^2^. Thus, JAK inhibition may improve ICB not only by antagonizing IFN-I signaling in CD8 T cells but also by inhibiting an IFN-driven resistant state in cancer cells.

## FIGURE LEGENDS

## SUPPLEMENTARY FIGURE LEGENDS

**Figure S1.**
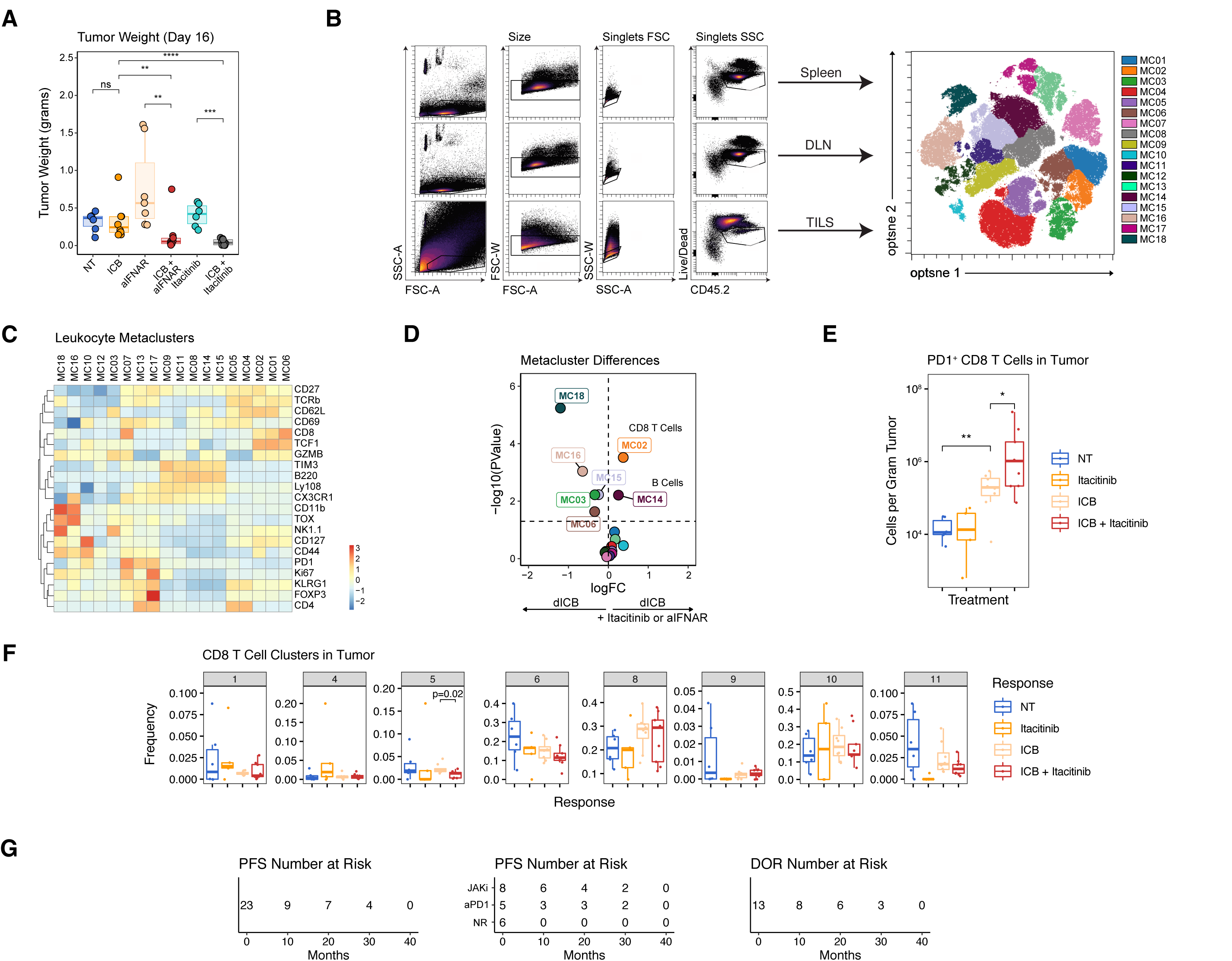
Tumor and immune response to anti-PD1 and a JAK1 inhibitor in mice and humans. **A)** Tumor weights from mice with Res 499 tumors at day 16 after the indicated treatment. **B-D)** Flow cytometry gating scheme of leukocytes from the indicated tissue and the generation of metaclusters using the indicated immune features **(B)**. The scaled MFI of these features for each metacluster is displayed in a heat map **(C)**. The significance of the differences in metacluster frequencies in the periphery (spleen) between mice treated with anti-PDL1 + anti-CTLA4 (ICB) versus ICB plus either itacitinib or anti-IFNAR1 are shown in the volcano plot **(D)**. **E-F)** Number of PD1^+^ CD8 T cells per gram of tumor for each of the indicated treatment groups **(E)**, and the frequency of different PD1^+^ CD8 T cell clusters in the tumor **(F)**. NT is non-treated. **G)** The number of patients at risk for the survival curves shown in Fig. 1J. PFS is progression-free survival and DOR is duration of response. For pairwise comparisons, a two-sided Wilcox test or t-test was used for non-parametric or parametric data, respectively. • p<0.1, * p<0.05, ** p<0.01, *** p<0.001.

**Figure S2.**
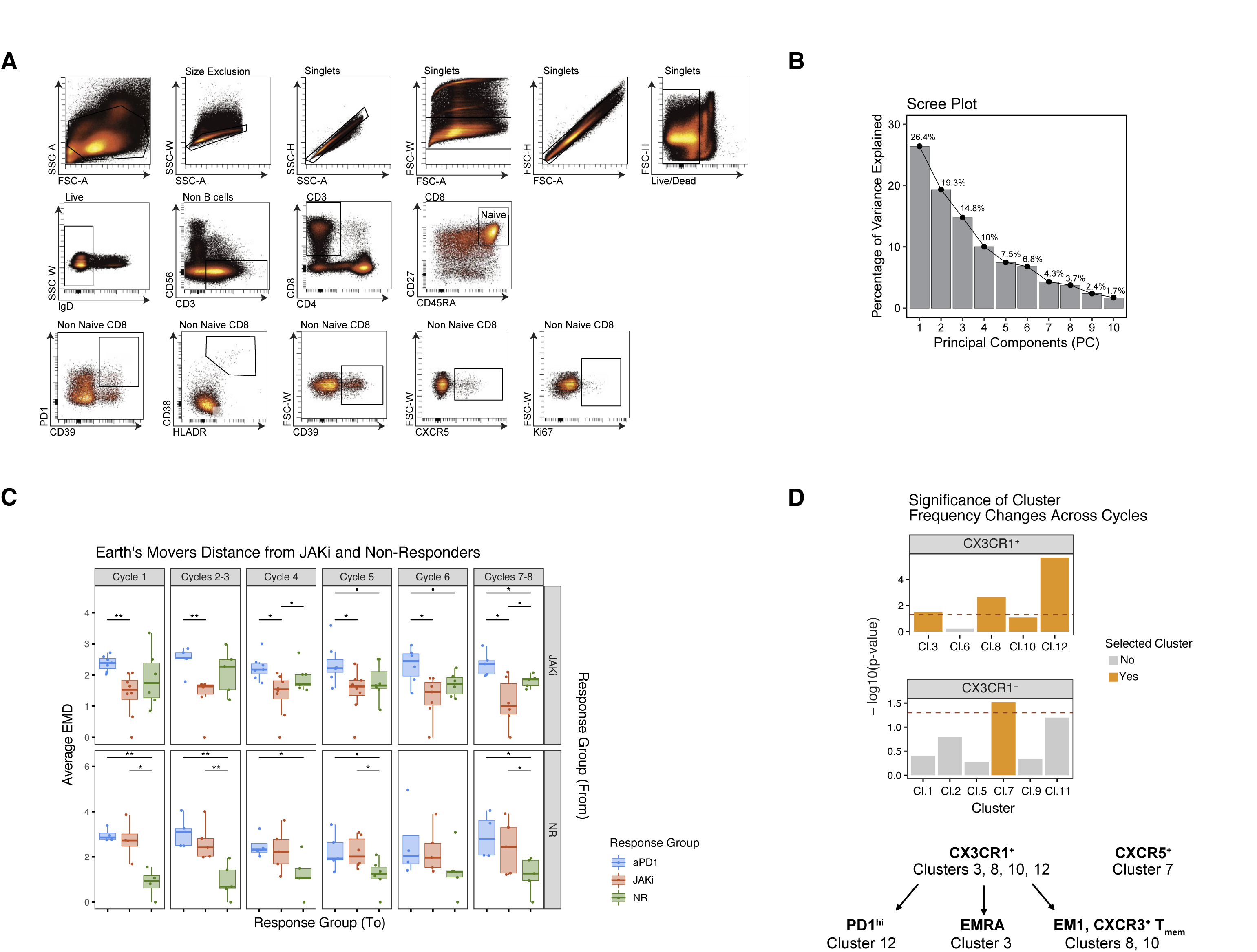
Features of CD8 T cell flow cytometry clusters that correlate with clinical response in NSCLC patients treated with pembrolizumab and delayed itacitinib. **A)** Flow cytometry gating scheme for non-naïve CD8 T cells from PBMCs from NSCLC patients prior to manual gating and principal components (PC) analysis or unbiased feature clustering. **B)** Scree plot from PC analysis showing percentage of variance explained for each PC. **C)** Average Earth Mover’s Distance (EMD) from CD8 T cell UMAPs (Fig. 2D) from JAKi responders (top row) or non-responders (bottom row) to the CD8 T cell UMAPs from each of the indicated response group (x-axis). Results are faceted by treatment cycles. **D)** Statistical significance (y-axis) for differences in individual CD8 T cell cluster frequencies (Fig. 2C) across treatment cycles. Results are displayed for CX3CR1^+^ clusters (top) and CX3CR1^−^ clusters (bottom). Those clusters with p-values less than or close to p=0.05 (doted line) were selected for further analysis (orange). A summary of the relationships between the selected clusters is shown below the bar plots. For pairwise comparisons, a two-sided Wilcox test or t-test was used for non-parametric or parametric data, respectively. • p<0.1, * p<0.05, ** p<0.01, *** p<0.001.

**Figure S3.**
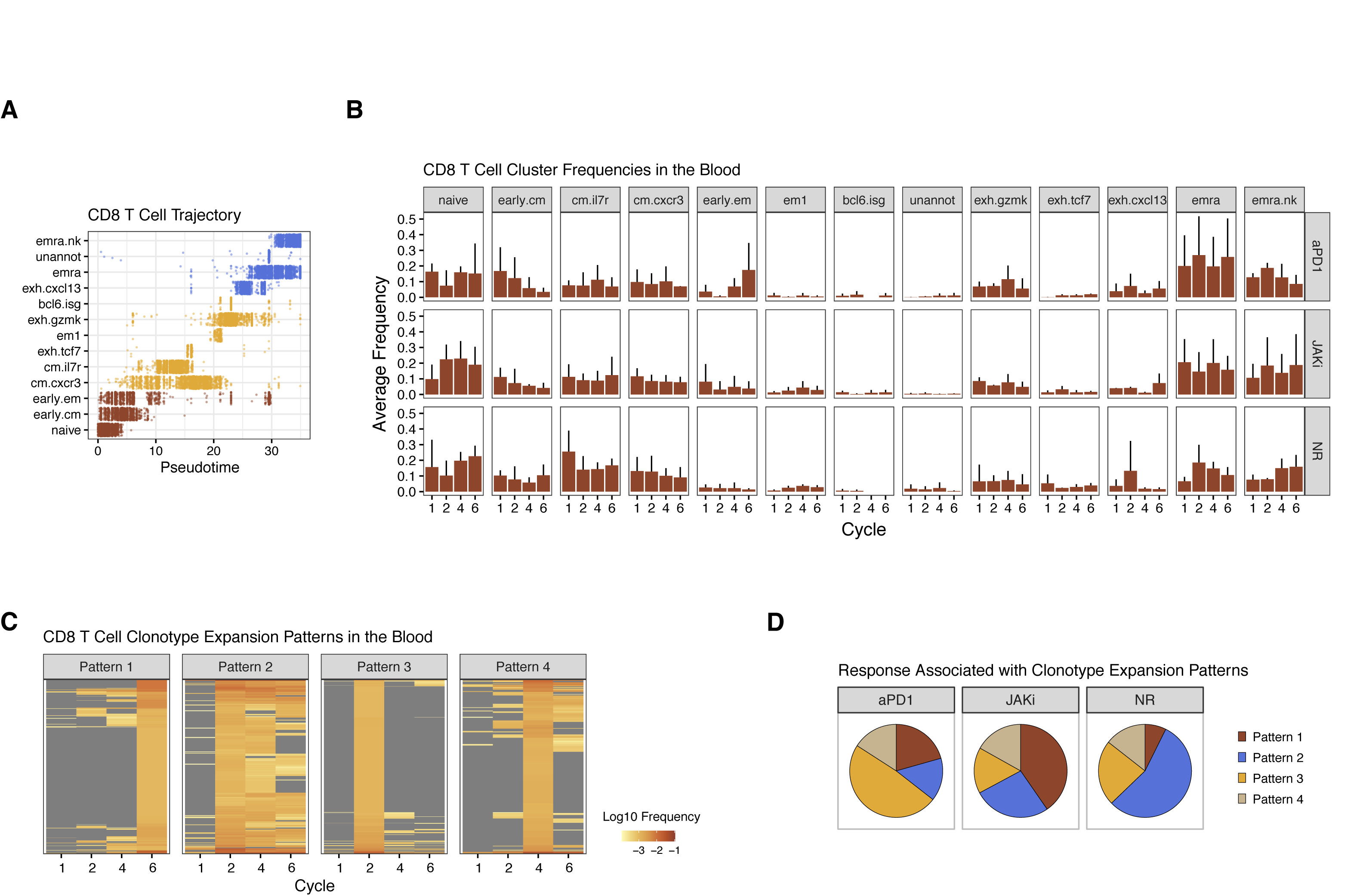
CD8 T cell differentiation, subset frequencies, and clonotype expansion patterns in NSCLC patients treated with pembrolizumab and delayed itacitinib. **A)** CD8 T cell differentiation trajectory analysis showing pseudotime for individual cells belonging to each of the indicated CD8 T cell subtypes (clusters) determined by single-cell RNA/TCR sequencing of CD8 T cells from NSCLC patients. **B)** Relative frequencies of CD8 T cell subtypes in the blood across treatment cycles. Shown are mean and standard deviations for patients belonging to each of the indicated clinical response groups. **C)** TCR clonotype expansion patterns from CD8 T cells in the blood determined by K-means clustering. For each heat map representing a distinct temporal pattern, rows are individual clonotypes selected based on filtering criteria to enrich for treatment-relevant clonotypes and color-coded by relative frequency. Grey colors indicate that the clonotype was below the frequency used for selection (see Methods). **D)** Pie charts showing the proportion of clonotypes from each expansion pattern associated with the indicated clinical response group.

**Figure S4.**
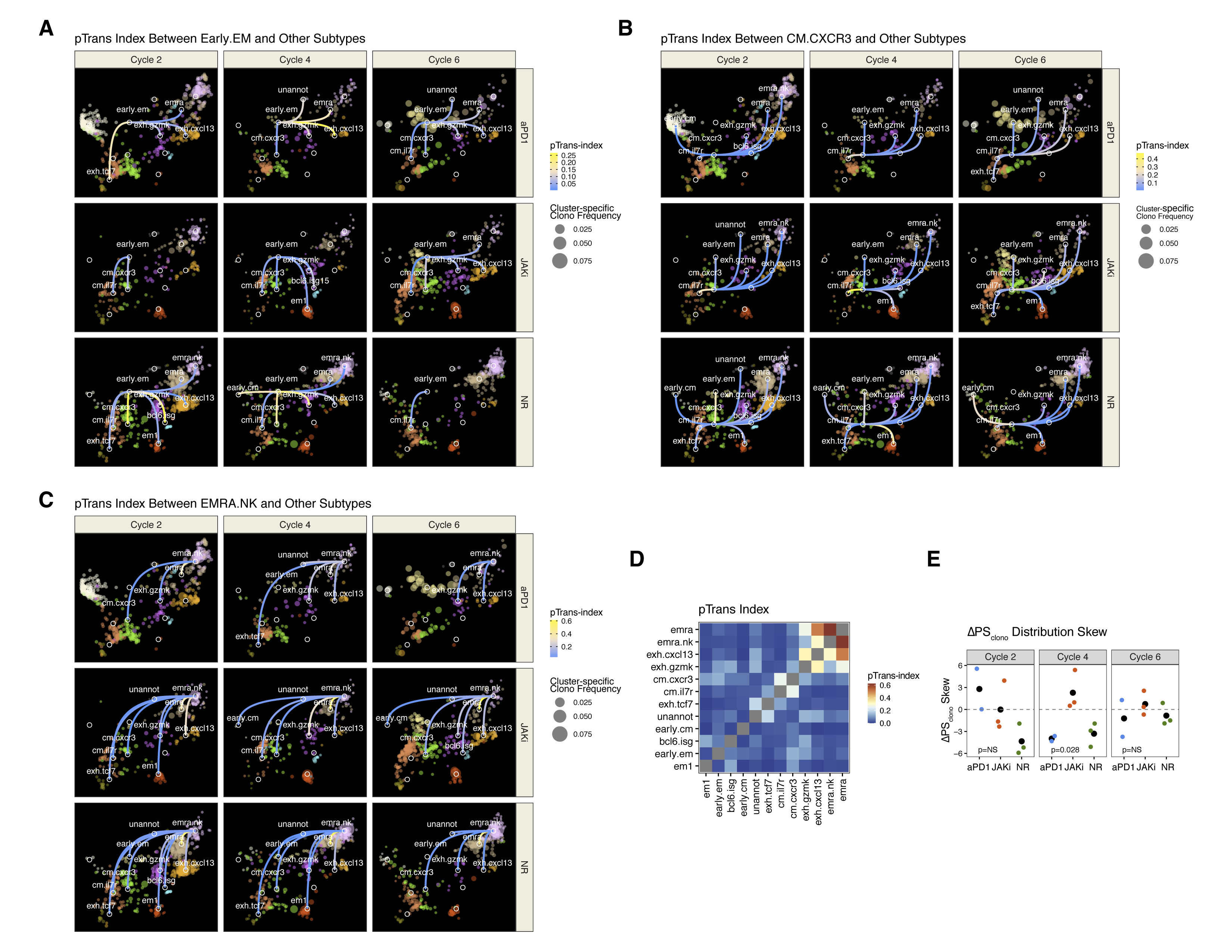
CD8 T cell differentiation, subset frequencies, and clonotype expansion patterns in NSCLC patients treated with pembrolizumab and delayed itacitinib. **A-C)** Blood-expanded CD8 T cell clonotypes belonging to the indicated subtype (color-coded) overlaid on the CD8 T cell UMAP (Fig. 3A) and faceted by treatment cycle and response group. The pairwise transition-index (pTrans-index) between the Early.EM subtype **(A)**, CM.CXCR3 subtype **(B)**, or the EMRA.NK subtype **(C)** and other subtypes are shown as lines between nodes and color-coded by the pTrans-index value (higher scores indicate greater TCR sharing). Subtype-specific clonotype frequency is represented by dot size. **D)** pTrans-index values between CD8 T cell subtypes. **E)** The skew of the ΔPS_clono_ distributions from each of the indicated response groups. Positive ΔPS_clono_ skew represents a distribution bias toward an increase in CD8 T cell clonotypes comprised of subtypes with high plasticity scores. For pairwise comparisons, a two-sided t-test was used. • p<0.1, * p<0.05, ** p<0.01, *** p<0.001.

**Figure S5.**
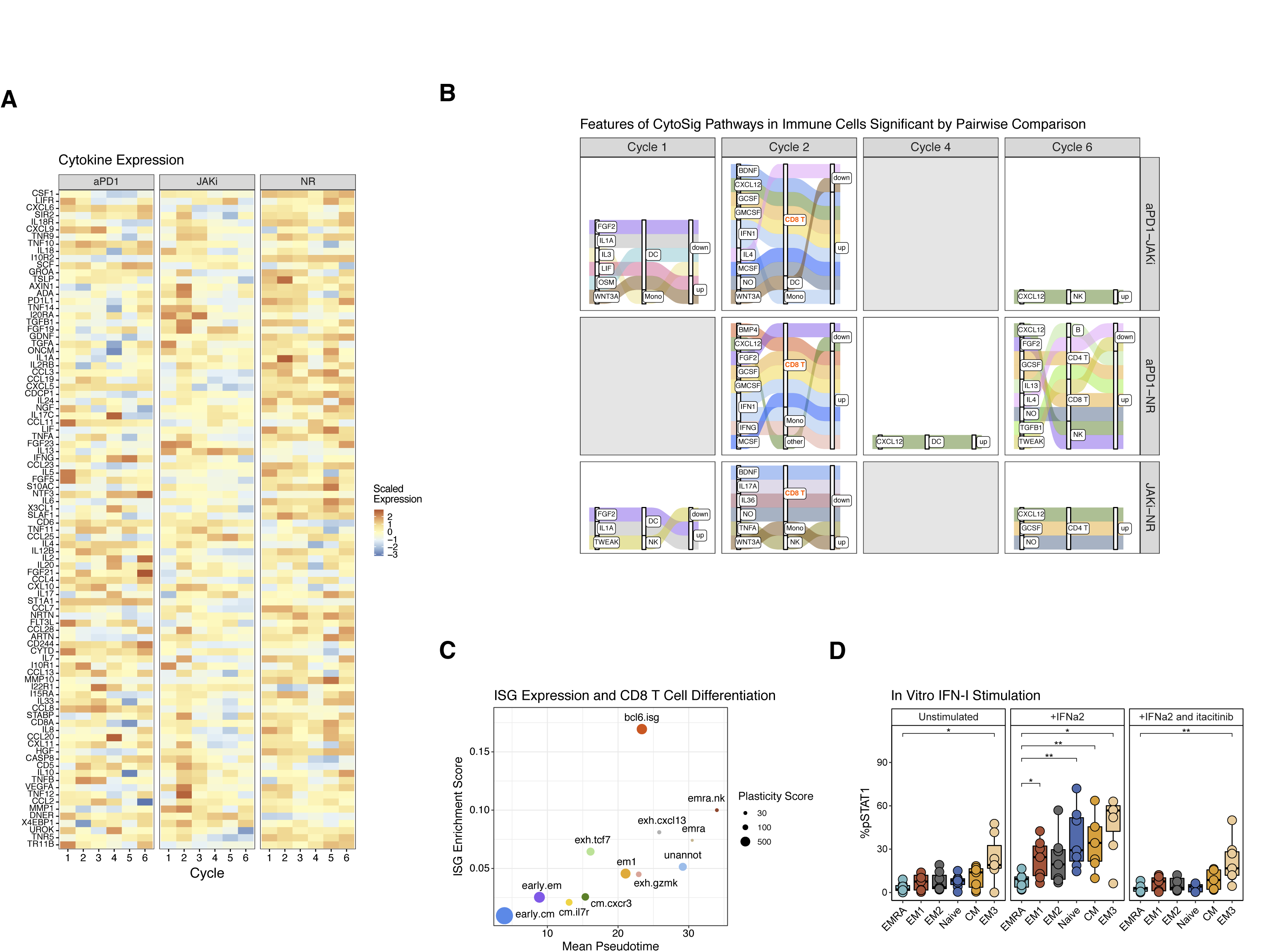
Serum proteins, cytokine signaling activity, and differentiation of immune cells from NSCLC patients treated with pembrolizumab and delayed itacitinib. **A)** Average normalized expression of serum proteins from NSCLC patients belonging to the indicated clinical response groups. **B)** Alluvial plot showing cytokine pathways from different immune cell type that significantly differ for each pairwise comparison between response groups (rows). The relationships between cytokine pathways, its immune cell type of origin, and whether activity is increased (up) or decreased (down) in the first group listed in the pairwise comparison are connected by ribbons. Results are faceted by treatment cycle. **C)** Relationship between ISG enrichment and average pseudotime from differentiation trajectory analysis of CD8 T cells belonging to each subtype. The plasticity score for each subtype is indicated by circle size. **D)** Percentage of CD8 T cells of the indicated subtype with phosphorylated STAT1 after *in vitro* stimulation with IFNA2 with or without itacitinib. For longitudinal cytokine pathway data, significance was determined by a repeated measures ANOVA using a mixed effect model and post-hoc interaction analysis. For pairwise comparisons, a two-sided t-test was used. • p<0.1, * p<0.05, ** p<0.01, *** p<0.001.

## MATERIALS AND METHODS

### Clinical trial

Patients with metastatic non-small cell lung cancer (NSCLC) who were treatment-naïve with PDL1 ≥50% by TPS PDL1 IHC 22C3 pharmDx assay (Dako North America), eastern cooperative group performance status (ECOG PS) 0-1, RECIST v 1.1 measurable disease amenable to biopsy, and no untreated brain metastases were enrolled (n=31 screened). Patients received pembrolizumab (200mg every 21 days) and Itacitinib 200mg daily by mouth was started Cycle 3 Day 1 of pembrolizumab and continued for 6 weeks. Of the 31 patients screened, 23 were enrolled between 1016/2018 and 3/4/2021 and received at least 1 cycle of pembrolizumab. Primary endpoints were: 1) overall response rate (ORR) determined by RECIST v1.1 partial (PR) and complete responses at 12 weeks 2) toxicity of pembrolizumab and itacitinib by common terminology criteria for adverse events (CTCAE) v5.0 (**Table S3**). Best ORR was defined as the best response determined by RECIST v1.1 over the study period. Secondary clinical objectives included progression free survival (PFS), and overall survival (OS) from initiation to study therapy until disease progression or death due to any cause, respectively. Duration of response (DOR) was defined as time from first response at 12-weeks until disease progression by RECIST v1.1. Response at 6-weeks was defined using a modified RECIST v1.1, allowing for changes of near -30% reduction to also be included as a PR. This resulted in a clear demarcation between responders (range: -27.3 to -61%) and non-responders (range: -19.5% to 150%). Patients were censored at the date of last follow up if they came off trial for reasons other than progression or death and at data cutoff (12/1/21) for those on trial without disease progression or death. Median PFS, DOR, OS were estimated using Kaplan Meier methodology. Paired blood and tissue samples were collected for several translational and exploratory objectives.

The clinical trial (ClinicalTrials.gov identifier: NCT03425006) was approved by the institutional review board at the University of Pennsylvania and was completed in accordance with international standards of good clinical practice. All patients provided written informed consent at the time of enrollment.

### Mice

Mice were maintained in a specific-pathogen-free facility at the University of Pennsylvania. Experiments and procedures were performed in accordance with the Institutional Animal Care and Use Committee (IACUC) of the University of Pennsylvania. Five-to seven-weeks-old female C57BL/6 mice were obtained from Charles River Production.

### Cell lines

Res 499 ICB-resistant B16-F10 melanoma was derived and cultured as previously described^11^. For ICB studies, 50,000 Res499 cells were mixed 1:1 with reduced growth factor basement membrane extract (BME type 2) and injected to the hind leg. Caliper measurements were started when palpable tumors were observed at day 11.

### In vivo mouse lymphocyte studies

Tumors, spleens, and draining lymph node (DLN) were harvested at day 16 post tumor implantation. For spleens and DLNs, single-cell suspensions were prepared after RBC lysis with ACK Lysis Buffer (Life Technologies). Tumors were weighed prior to enzymatic digestion with Type 4 collagenase and DNAse 1 at 1mg/mL. After enzymatic digestion or ACK Lysis, all tissues were filtered through 100 micron filters. Cells were stained with Fc Block and Zombie Live/Dead stain for 10 minutes prior to surface stain. Surface stain was done for 30 minutes at room temperature. Samples were fixed and permeabilized by incubating in 100 μl of Fix/Perm buffer at room temperature for 30 minutes and washed in Perm Buffer. Intracellular stains were performed overnight at 4C. Cell counting beads were spiked into each sample prior to data acquisition. Data acquisition was done on a FACSymphony™ A5 and analyzed using OMIQ and/or R. See Resource Table for list of antibodies and buffers.

### Human antibody panels and staining

Approximately 1×10^6^ to 5×10^6^ frozen PBMCs were used per patient per timepoint. PBMCs were thawed into 10% complete RPMI with DNase. Cells were stained with live/dead and Fc block for 10 minutes at room temperature. Chemokine receptors were stained at 37C for 20 minutes followed immediately by surface stain for 30 minutes at room temperature. Samples were fixed and permeabilized by incubating in 100 μl of Fix/Perm buffer at room temperature for 30 minutes and washed in Perm Buffer. Intracellular stains were performed overnight at 4C. Data acquisition was done on a FACSymphony™ A5 and analyzed using OMIQ and/or R. See Resource Table for list of antibodies and buffers.

### Flow cytometry feature clustering

For quantification and statistical analysis of flow cytometry data, the *flowCore* and *flowWorkspace* R packages and custom R analysis pipelines were used. A downsampled feature matrix was then created by equal random sampling of cells from each FCS file. This downsampled data were then used for dimensionality reduction by UMAP, as implemented in the *umap* R package, and clustering by self-organizing maps, as implemented in the *FlowSOM* R package. The number of clusters (K) was determined using random forest with stratified sampling, as implemented in the *randomForestSRC* R package, to estimate an out-of-bag overall prediction error rate for a range of K values. Then, a value for K was selected to maximize the number of clusters without a steep increase in error rate. Using this reference map, a random forest classifier was then trained and used to predict cluster membership for all cells for all samples. To determine similarity of UMAPs for samples belonging to different groups, the *transport* R package was used to calculate an Earth Mover’s Distance for each sample compared to all others.

### Olink

Cytokines were measured from EDTA-plasma using the Olink Extension Assay (PEA) to measure 92 unique analytes. In brief, oligonucleotide labeled capture antibodies bind target cytokines and subsequently hybridize. The hybridized product was then amplified and measured by qPCR, allowing for parallel detection of 92 cytokines within the same sample.

### pSTAT1 detection

Healthy donor PBMCs were plated overnight in RPMI at 1×10^6^ cells per 100uL. The next day, PBMCs were stimulated with 20ng/mL IFNa2 (Biolegend #592704) for 15 minutes. Reactions were stopped with a final concentration of 2% PFA and kept in methanol O/N in -80C. The next day, cells were washed with PBS and stained with a pSTAT detection panel for 1 hour at room temperature. Data acquisition was done on a FACSymphony™ A5 and analyzed using OMIQ.

### Single-cell RNA and TCR sequencing and processing

PBMCs from select patients were sorted for total live cells or live CD8^+^ cells on a BD FACs Aria II. Cells were sorted into GEMs using 10x Chromium Controller and were made into libraries following the Chromium Next GEM Single Cell 5’ Reagent Kits v2 (Dual Index) Protocol. Libraries were sequenced using a NovaSeq 6000. Sequencing data was processed using *CellRanger* pipeline v5 (10x Genomics). BCL files were converted to FASTQ and aligned to the human genome (GRCh38) to generate count matrices. For TCR libraries, BCL files were converted to FASTQ and CellRanger VDJ pipeline was used for sequence assembly and clonotype calling. Cells with >10% mitochondrial DNA, <200 or >2500 RNA features were filtered out and conditions integrated using *Seurat* v3.2.0. UMI barcodes were used to combine cell expression data with clonotype data.

### Human CD8 T cell reference mapping and annotation

Using the Seurat objects for scRNA/TCR-sequencing data from patient PBMC-sorted CD8 T cells from all study timepoints (start of cycles 1, 2, 4, 6), 1000 cells with a rearranged TCR were randomly sampled and a separate Seurat objected was created. Processing to filter out cells with high mitochondrial DNA and variations in UMI count were carried out as described. Data were then log normalized, variable features identified, scaled, and integrated using the RPCA reduction method from *Seurat* with k.anchors = 20. Predicted cell doublets were identified with the *DoubletFinder* R package. After dimensionality reduction by PCA and UMAP, cell clustering was performed by shared nearest-neighbor. Very sparse clusters or clusters consisted primarily of predicted doublets were removed along with any remaining predicted cell doublets. This resulted in a CD8 T cell reference consisting of approximately 20,000 cells. After re-clustering, the reference was annotated using marker gene expression and enrichment of CD8 T cell subtype gensets by GSVA (described below). This annotated CD8 T cell reference was then used to map 1) scRNA/TCR-seq data of CD8 T cells sorted from PBMCs, and 2) CD8 T cells from sequenced PBMCs from the same sample. For the CD8 T cells from scRNA/TCR-sequenced PBMCs, the PBMC data were first mapped to the Azimuth Human PBMC reference using MapQuery from *Seurat*. CD8 T cells identified from the level one predicted cell type annotation were subsetted to create a separate Seurat object that was then merged with the Seurat object of sorted CD8 T cells from the same sample (i.e., same patient and same timepoint). The merged CD8 T cell data were then mapped to the CD8 T cell reference using MapQuery.

### Gene set enrichment and scores

Gene sets for human CD8 T cells from PBMCs and from a human pan-cancer T cell atlas were used to aid in cluster annotation and analysis^9,21^. For the human PBMC CD8 T cell gene sets, the top 250 genes from each set were used. For the CD8 T cell gene sets from the human pan-cancer T cell atlas, the top 100 genes or all the genes in the gene set were used, whichever was smaller. The scRNA-sequencing data from all cells in a cluster were averaged to create pseudo-bulk data. Then, an enrichment score for each gene set of interest was determined using GSVA. To determine gene set scores for individual cells, the AddModuleScore function of *Seurat* was used.

### Pseudotime trajectory analysis

Pseudotime trajectory analysis was carried out using *Monocle3*. The integrated gene expression data from the CD8 T cell reference map was used as ordering genes to construct pseudotime trajectories. The naïve CD8 T cell cluster was selected as a starting root state. Assigned pseudotime values for each cell was then used in downstream analysis.

### CD8 T cell clonotype filtering and analysis

To enrich for well-annotated and treatment-relevant CD8 T cells and clonotypes, a set of filtering criteria were applied to each sample. These criteria included only keeping CD8 T cells with: 1) a single rearranged TCR comprised of a single TCR alpha and TCR beta chain, 2) predicted singlet by *DoubletFinder*, and 3) reference mapping annotation score > 0.50. CD8 T cells that met these criteria were then assigned to a clonotype using the amino acid sequences of the CDR3 region of TCR alpha and beta. Then, filtering criteria for clonotypes were used to keep only assigned clonotypes that: 1) was not exclusive to naïve CD8 T cells, 2) had a relative frequency in the blood of at least 0.001, and 3) increased in frequency above baseline by at least 5-fold. To examine clonotypes shared by the tumor and the blood, the CDR3 of the TCR beta chain from Adaptive TCR-sequencing was matched to the TCR beta chain from 10X Genomics sequencing with no requirement for a minimum frequency in the blood or increase above baseline.

### TCR sharing and clonotype plasticity

Assessment of clonotype sharing between two CD8 T cell states was calculated using *Startrac*^24^ to determine the pairwise transition index (pTrans-index). The plasticity score (PS) for each CD8 T cell subtype was then defined as the reciprocal of the variance of its pTrans-index with each of the other subtypes. A clonotype plasticity score (PS_clono_) was defined as the average of the plasticity scores for all CD8 T cells subtypes comprising the clonotype. The ΔPS_clono_ is the difference between the PS_clono_ at a given treatment cycle from the PS_clono_ at baseline (start of cycle 1).

### Cytokine signaling activity score

The cytokine signaling activity of immune cells from scRNA-sequencing data was predicted using *CytoSig*^33^. The scRNA-sequencing data from all cells in a cluster were averaged to create pseudo-bulk data. This was then used with the *CytoSig* Python package to return the predicted cytokine signaling activity scores for different immune cells or CD8 T cell subtypes.

### Statistical analysis

The significance of changes across treatment cycle for CD8 T cell frequencies (flow cytometry), clonotype frequencies (scRNA/TCR-seq), serum protein levels (Olink), and cytokine signaling activity scores (CytoSig) were examined by a repeated measures ANOVA using a mixed effect model implemented in the *nlme* R package. If the main effect was significant, post-hoc interaction analysis was used to determine within-group or between-group differences using the *phia* R package. Z-scores, scaled, or normalized values were used for the models.

Differences in mouse tumor growth were determined using a mixed-effect regression model set to a log-normal distribution using the *MASS* R package. For survival analysis, the Kaplan-Meier estimate and a log-rank test from the *survival* R package were used. Median follow-up times were calculated using the *prodlim* R package and the reverse Kaplan-Meier method. For simple two group comparisons, a two-sided Wilcoxon test or t-test was used for non-parametric or parametric data, respectively. For multiple groups, a Kruskall-Wallis or ANOVA test was used along with TukeyHSD for post-hoc testing. Normality was assessed using a Shapiro’s test. The *FactoMineR* R package was used for principal components analysis.

## Data Availability

All data produced in the present study will be made available upon publication of the manuscript in a peer-reviewed journal.

---

**Table S1.** Baseline Patient Characteristics

|  | All Patients |
| --- | --- |
| **Number of Patients** | 21 |
| **Age - Mean (Std)** | 62.95 (8.54) |
| **Sex - Number (%)** |  |
| Male | 10 (47.6) |
| Female | 11 (52.3%) |
| **PDL1 - Mean (Std)\*** | >77% (>15.3%) |
| **Smoking - Number (%)\*** |  |
| Current | 10 (50%) |
| Former | 8 (40%) |
| Never | 2 (10%) |
| **Histology - Number (%)\*** |  |
| Adenocarcinoma | 15 (75%) |
| Squamous cell | 5 (25%) |
| **Race - Number (%)\*** |  |
| Black | 6 (30%) |
| White | 12 (60%) |
| Unknwn | 2 (10%) |

\* Missing patient value

**Table S2.**
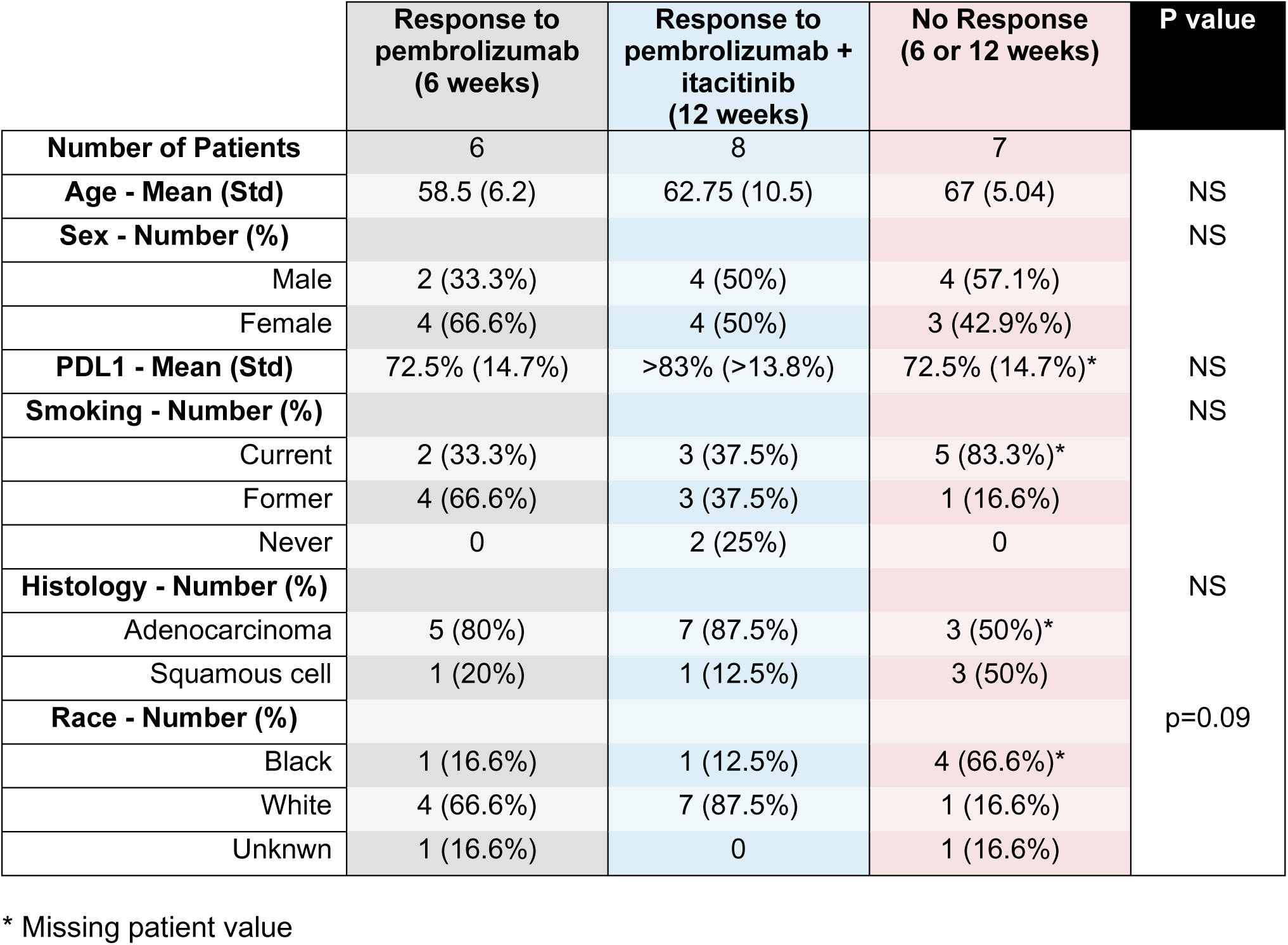
Characteristics of Patient Response Groups

**Table S3.** Adverse Events Related to Study Drug

| Category | Pembrolizumab |  | Itacinitib |  |
| --- | --- | --- | --- | --- |
|  | Grade 1-2 | Grade 3+ | Grade 1-2 | Grade 3+ |
| Gastrointestinal disorder | 1 | 2 | 0 | 0 |
| Blood and lymphatic disorder | 0 | 0 | 1 (anemia) | 0 |
| Endocrine disorder | 2 | 0 | 0 | 0 |
| Electrolyte disorder | 1 | 1 | 0 | 0 |
| Musculoskeletal disorder | 4 (arthralgia) | 0 | 0 | 0 |
| Nervous System disorder | 1 | 2 | 2 | 0 |
| Pneumonitis | 1 | 1 | 0 | 0 |
| Skin disorders | 0 | 0 | 1 | 0 |

